# Unveiling Shared Genetic Links between Blood Cell Traits and Cardiovascular Diseases

**DOI:** 10.1101/2024.10.23.24315926

**Authors:** Luke Kong, Kaixin Yao, LeiLei Zheng, Yuhui Zhao, Miaoran Chen, Ziyi Han, Qian Wang, Qifan Feng, Yongqiang Lv, Biyun Zhang, Feng Li

## Abstract

Previous studies have linked blood cell traits (BCTs) to cardiovascular diseases (CVDs) risks, but the common genetic mechanisms underlying heritable phenotypes remain unclear. Our study used multiple analytical approaches including single nucleotide polymorphisms, genes, pathways, and protein targets to reveal common genetic elements. We confirmed both genome-wide and local genetic associations between BCTs and CVDs, identifying key pleiotropic loci and genes contributing to these links. Specifically, *ALDH2*, *MAPKAPK5*, and *ACAD10*, all located at 12q24.1, are associated with leukocyte-CVD traits. *TNFSF12* at 17p13.1 and *ABO* at 9q34.2 correlate with platelet-CVD traits, while *ZNF664* and *CCDC92*, also at 12q24.1, are linked to erythrocyte-CVD traits. Our findings also highlight multiple key trait-specific pathways mediating these phenotypic associations and potential therapeutic targets that may inform future clinical interventions. These insights significantly advance our understanding of the genetic interplay between BCTs and CVDs, underscoring the importance of focusing on BCTs to prevent cardiovascular conditions.

**Highlights:**

- There were wide genetic correlation and overlap between BCT and CVD.
- Key pleiotropic loci and genes for three BCT-CVD trait pairs were identified.
- Key signature-specific pathways mediating the BCTs-CVDs association were identified.
- Potential therapeutic targets for BCTs-CVDs were identified.

**eTOC blurb:** Kong et al. report extensive genetic correlations and genetic overlaps between blood cell traits (BCTs) and cardiovascular diseases (CVDs), revealing key pleiotropic loci and genes contributing to these associations, as well as multiple trait-specific pathways mediating these phenotypic associations. Potential therapeutic targets to inform future clinical interventions were also identified, laying the foundation for understanding the genetic interactions between BCTs and CVDs.

**Graphical abstract:** 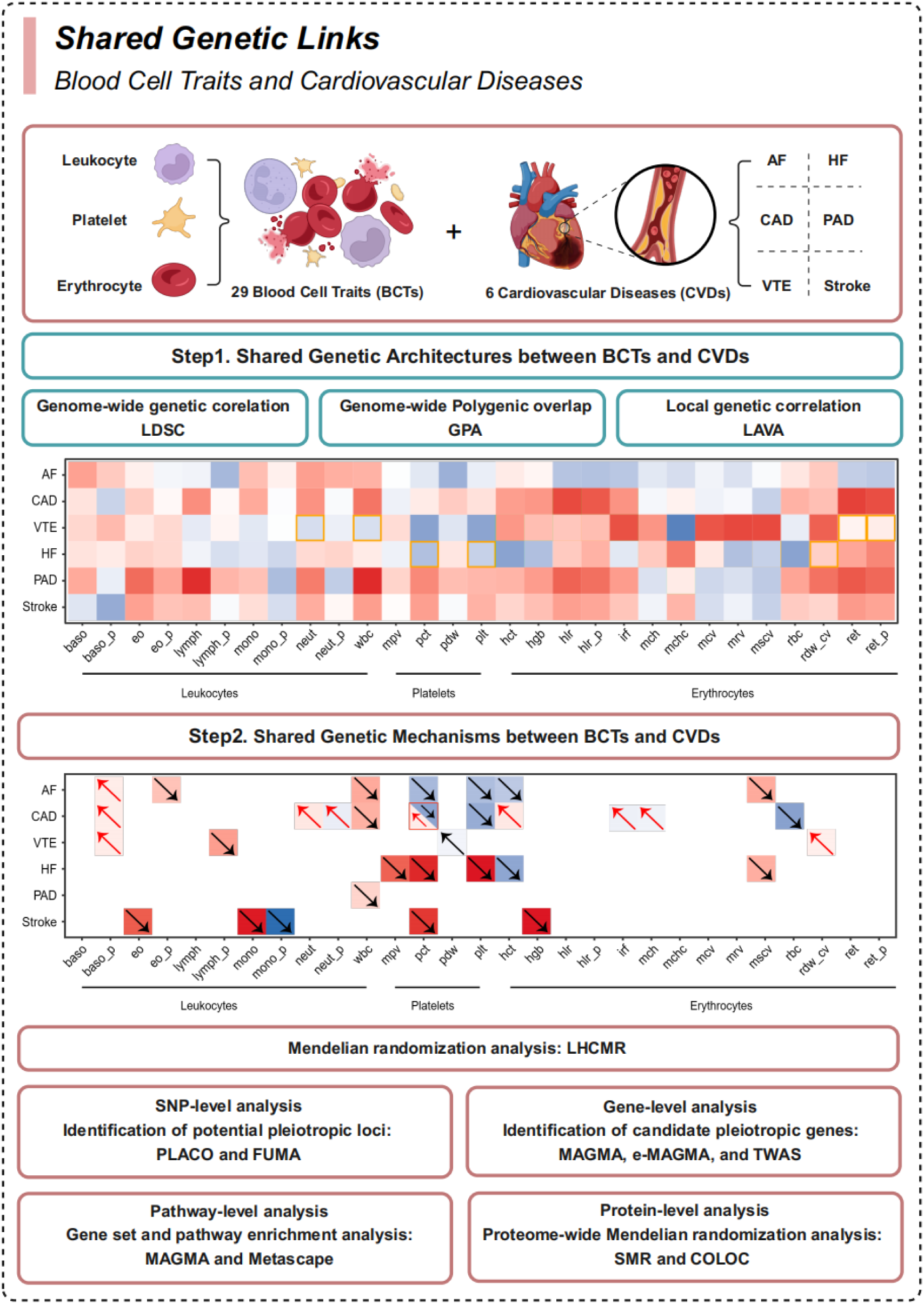

## Introdution

Cardiovascular diseases (CVDs), which encompass a range of conditions affecting the heart and blood vessels, are the leading cause of death and disability worldwide^1,2^. A primary contributor to CVDs, atherosclerosis, is significantly influenced by various blood cell traits (BCTs)^3,4^. For example, erythrocytes, critical in maintaining physiological hemodynamics, can exacerbate arterial wall pathologies and, through interactions with immune cells, potentially accelerate atherosclerosis due to oxidative stress^5^. Similarly, the secretory functions of leukocytes, especially lymphocytes, are pivotal as they release proinflammatory cytokines and proteases that may lead to plaque rupture^6^. Platelets also play a key role in hemostasis and thrombosis and enhancing the inflammatory environment, thus promoting the recruitment of inflammatory cells to lesions and releasing inflammatory mediators^7^. Extensive observational research has consistently demonstrated a robust association between alterations in BCTs and increased risk of CVDs. Recent studies have identified erythrocyte count (RBC), hematocrit (HCT), mean corpuscular volume (MCV), and red cell distribution width (RDW-CV) as potential biomarkers for cardiovascular risk.^8^. Additionally, conditions characteristic of CVDs can prompt the bone marrow to release immature cells or increase other cell populations, depending on the severity. Notably, the leukocyte is a cost-effective, widely used diagnostic tool in clinical practice, with insights from the Framingham Heart Study highlighting its role as an indicator of elevated CVD risks^9^. More precise predictions of CVD risk come from specific leukocyte subtypes, such as monocytes^10^, lymphocytes^11^, and neutrophils^12^, rather than total leukocyte count (WBC) alone. Furthermore, studies have shown that platelet metrics like platelet component distribution width (PDW), mean platelet volume (MPV), and platelet count (PLT) are critical indicators of CVD risk^13^. These insights underscore the importance of monitoring alterations in blood cell functions for effective diagnosis, risk stratification, and predicting outcomes in CVDs.

Epidemiological studies have consistently shown that BCTs are closely associated with CVDs, with this relationship likely influenced by shared genetic factors^14,15^. Genome-wide association studies (GWAS) have identified numerous genetic variants common to both BCTs and CVDs, such as *SH2B3* and *HFE*, which are associated with RBC and coronary artery disease (CAD), respectively^16–18^. This suggests that pleiotropy, the influence of a single genetic variant on multiple traits, may be fundamental to understanding the genetic basis of these complex traits. Pleiotropy manifests in two primary forms: vertical and horizontal^19^. Vertical pleiotropy occurs when single nucleotide polymorphisms (SNPs) influence one trait, which subsequently affects another, permitting the use of Mendelian randomization (MR) to estimate causal relationships between traits. For example, Hashfield *et al.* demonstrated a positive causal relationship between PLT and eosinophil percentage of leukocytes (EO_P) with an increased risk of ischemic stroke and its subtypes^20^. Similarly, genetic predispositions towards high RBC and low monocyte count (MONO) correlate with increased venous thromboembolism (VTE) risk^21^. However, existing MR studies have yet to explore these trait categories fully. On the other hand, horizontal pleiotropy occurs when a genetic variant independently affects multiple phenotypes or influences intermediate processes between these phenotypes. Recent advances in statistical tools for genomics have underscored the importance of horizontal pleiotropy in elucidating the shared genetic basis of complex traits. For example, Yang *et al.* reported an extensive landscape of genetic correlations between 29 BCTs and 11 neurological and psychiatric diseases^22^. Despite these insights, previous studies have yet to comprehensively explore the relationship between BCTs and CVDs. The complex association patterns and underlying mechanisms of their connection remain largely uncharted. Therefore, systematic analyses are necessary to determine whether shared genetic architectures and molecular pathways exist between BCTs and CVDs, potentially unveiling new insights into their biological mechanisms.

In this study, we comprehensively analyzed the latest and most extensive GWAS summary data from individuals of European ancestry to uncover the shared genetic architecture and mechanisms underlying 29 BCTs and six major CVDs (Table 1). Our initial analyses focused on identifying the shared genetic structure between BCTs and CVDs, utilizing methods to analyze genetic correlations and overlaps. Then, employing MR analysis within a framework of vertical pleiotropy, we explored evidence for causal relationships between BCTs and CVDs. This groundwork facilitated a cross-trait analysis, pinpointing pleiotropic SNPs at the SNP level, and allowed us to identify candidate pleiotropic genes through positional mapping and expression quantitative trait loci (eQTL) mapping. Further investigations included an enrichment analysis of biological pathways and the identification of pathogenic plasma proteins, assessing their potential as therapeutic targets. This multifaceted approach has illuminated complex genetic networks linking hematological markers to cardiovascular health, offering insights that could pave the way for novel diagnostic and therapeutic strategies.

**Table 1:**
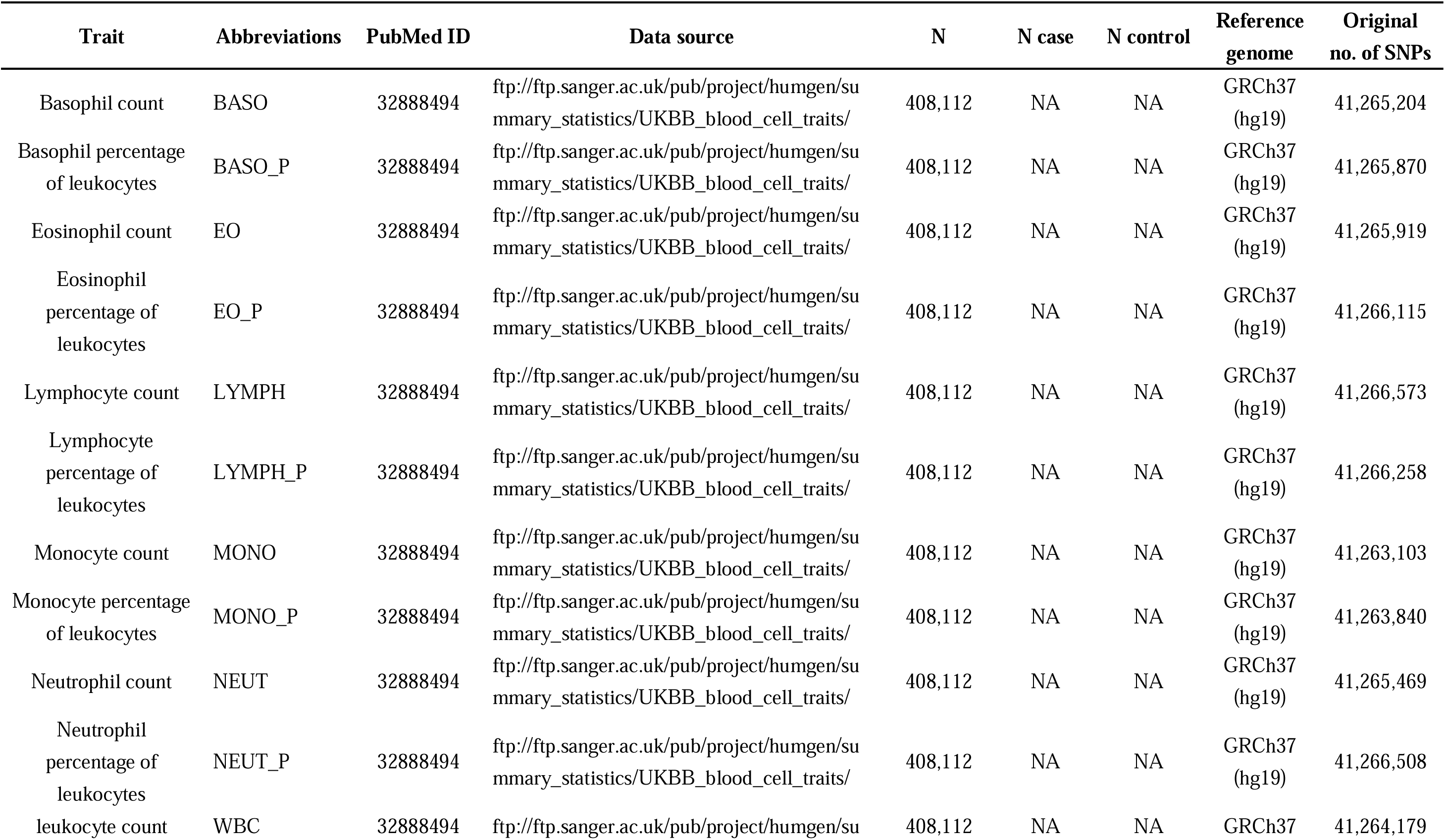
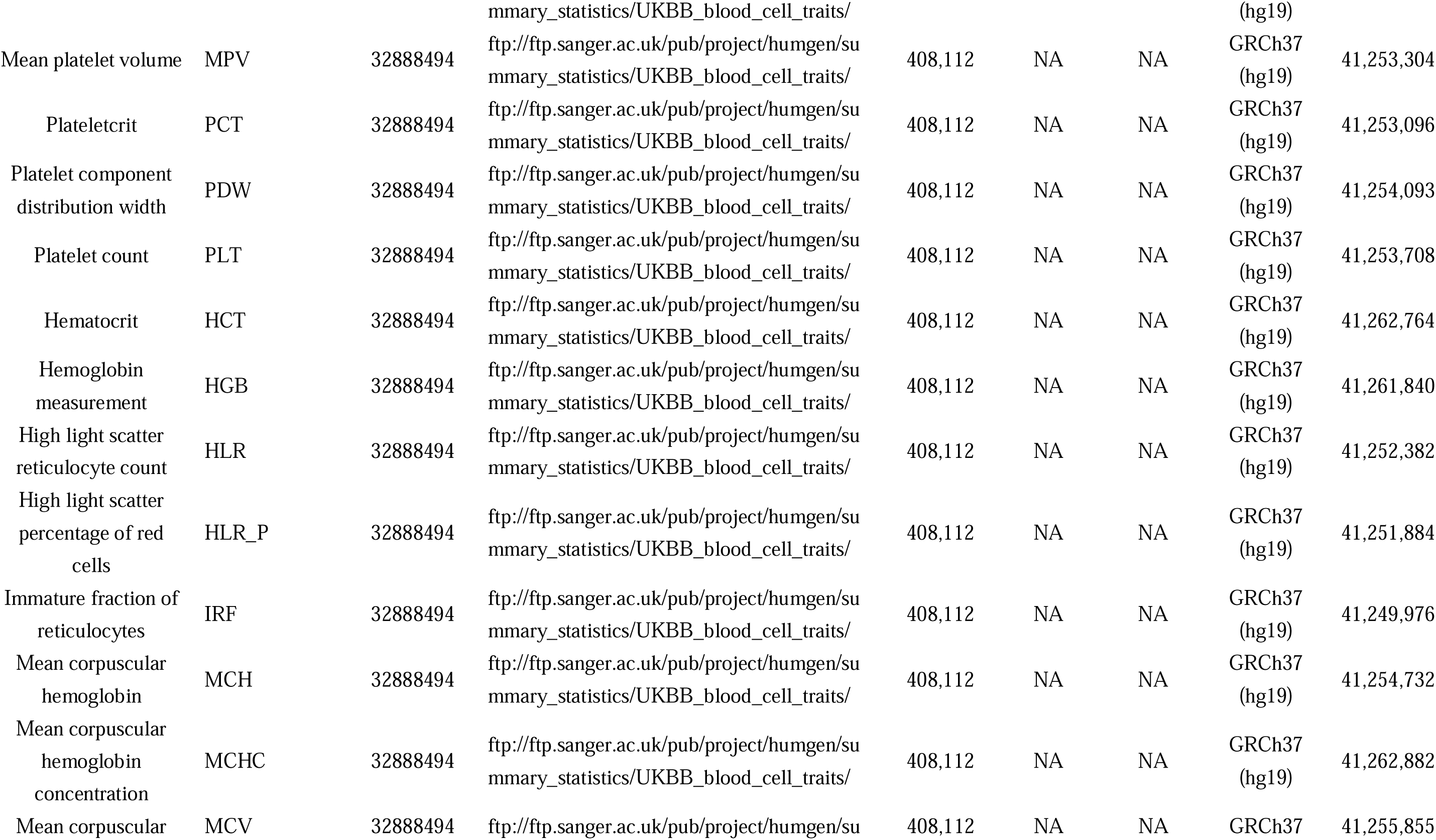
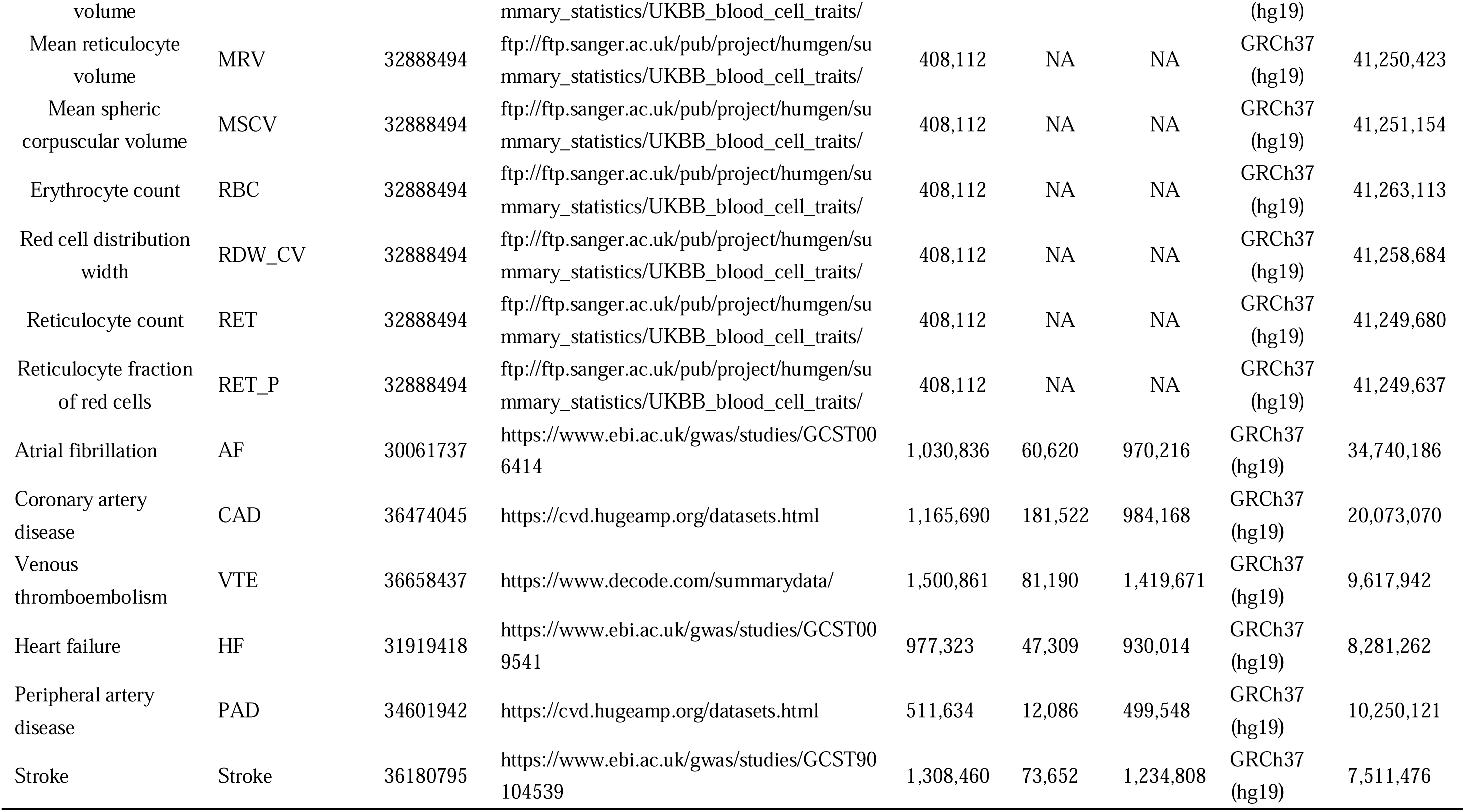
Overview of 29 blood cell traits and 6 cardiovascular diseases included in this study. Note: Overview of 29 blood cell traits and 6 cardiovascular diseases, abbreviations as used throughout the manuscript, associated PubMed ID, Data source and Year of publication, the sample size, population and reference genome on which summary statistics are based, and the number of SNPs included in the original summary statistics, before we applied filtering.

## Result

### Genome-wide genetic correlation between BCTs and CVDs

We assessed the SNP-based heritability (*h^2^_SNP_*) of 29 BCTs and 6 major CVDs using linkage disequilibrium (LD) score regression (LDSC) and performed genome-wide genetic correlation (*r_g_*) analysis of these 174 trait pairs. Univariate LDSC analysis showed that *h^2^_SNP_* estimates for BCTs were consistently higher than those for CVDs, ranging from 5.2% to 34.9%, with the highest estimate for MPV (*h^2^_SNP_* = 34.88%, SE = 0.036). In contrast, *h^2^_SNP_* estimates for CVDs were significantly lower, all below 5%, with the smallest estimate for Stroke (*h^2^_SNP_* = 0.60%, SE = 0.0005) and the highest estimate for CAD (*h^2^_SNP_* = 3.25%, SE = 0.0019) (Supplementary Table 1). Subsequent bivariate LDSC analysis found nominally significant associations for 36 (20.3%) of 174 trait pairs (*P* < 0.05). Of these, 26 trait pairs showed positive *r_g_*, especially between WBC and PAD, with a *r_g_* of 0.115 (SE = 0.053). In contrast, the most significant negative *r_g_* was between mean corpuscular hemoglobin concentration (MCHC) and VTE, with a *r_g_* of -0.101 (SE = 0.028). Using a less stringent 5% false discovery rate (FDR) threshold, we found that three blood cell count-related tra<u>it</u>s, including platelet crit (PCT), PLT, and MCHC, were associated with a lower risk of VTE. Six different BCTs (neutrophil count (NEUT), WBC, high light scatter reticulocyte count (HLR), high light scatter percentage of red cells (HLR_P), reticulocyte count (RET), and reticulocyte fraction of red cells (RET_P)) were significantly associated with an increased risk of CAD. In contrast, six different BCTs (immature fraction of reticulocytes (IRF), mean corpuscular hemoglobin (MCH), MCV, mean reticulocyte volume (MRV), mean spheric corpuscular volume (MSCV), and red cell distribution width (RDW_CV) were significantly associated with an increased risk of VTE. Notably, four trait pairs previously identified as having the same direction of genetic correlations (including HLR-CAD, MRV-VTE, MCV-VTE, and RDW_CV-VTE) confirmed significant correlations between them^21,23–26^ (Fig. 1 and Supplementary Table 2).

**Figure 1:**
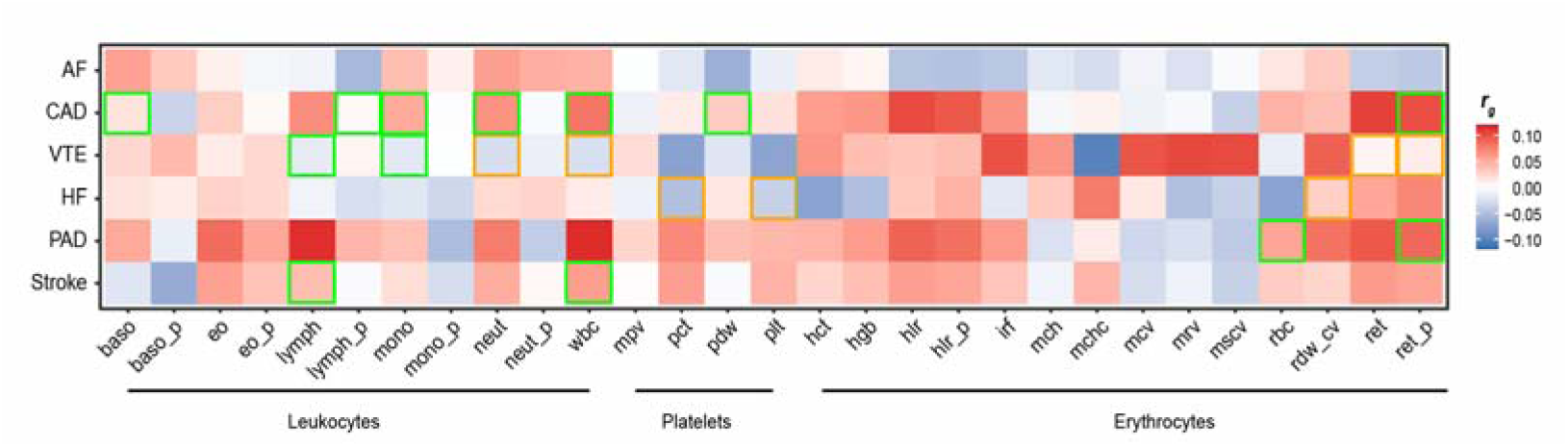
Estimated genome-wide genetic correlations between BCTs and CVDs using LDSC. Genome-wide genetic associations between BCTs and CVDs. Orange or green borders highlight significant genetic correlations (with estimates provided) if they were FDR significant (FDR < 0.05) or nominally significant (p < 0.05), respectively.

### Local genetic correlation between BCTs and CVDs

LDSC is used to estimate additive genetic effects on traits across the genome, however, when genetic signals show opposite correlations in different genomic regions, the cumulative genome-wide *r_g_* can asymptotically approach zero. To address this issue, we used Local Analysis of [co] Variant Annotation (LAVA) to perform more detailed analyses within shorter genomic regions, specifically focusing on local genetic correlations (local-*r_g_s*) between 29 BCTs and 6 CVDs. Applying it to 80,126 regions, we identified 28,887 univariate genetic signals associated with BCTs in 2,219 unique regions and 2,716 signals associated with CVDs in 1,472 unique regions, all with p-values less than 1×10^-4^ (Supplementary Table 3). Subsequent bivariate analysis revealed significant local-*r_g_s* (FDR < 0.05) for all trait pairs, covering 5,233 genetic regions (including 898 unique regions)(Supplementary Fig.1 and Supplementary Table 4). Interestingly, there is mixed local genetic effects on trait pairs where genome-wide *r_g_* is not significant. For example, between MSCV and CAD, 41.4% of the partitions showed positive associations, 59.6% showed negative associations, suggesting that genome-wide *r_g_* may underestimate the polygenic overlap of these trait pairs. Notably, LD block 1841 on chromosome 12 was associated with more than half of the trait pairs, followed by LD block 1479 on chromosome 9 and LD block 1480 on chromosome 12, which showed correlations with 68 and 47 trait pairs, respectively. Then, Hypothesis Prioritisation for multi-trait Colocalization (HyPrColoc) further identified the common causal variants among them and indicated a high association frequency for pleiotropic SNPs in the 12q24.12 region. Notably, rs3184504 (mapped to the *SH2B3* gene) showed strong evidence of colocalization between 19 BCTs and 2 CVDs (CAD and VTE), with a posterior probability (PP) > 0.7. In addition, rs4378452 (mapped to the *CUX2* gene) was identified as a potential common causal variable between 22 BCTs and CAD.

### Extensive genetic overlap between BCTs and CVDs

Consider that *r_g_* can not characterize detailed association patterns at individual loci, and nonsignificant estimates do not necessarily indicate the absence of a common genetic background. Therefore, we further applied genetic analysis combining pleiotropy and annotation (GPA) approach to explore the overlapping genetic variants of 29 BCTs and 6 CVDs, enhancing the understanding of the common genetic landscape of BCTs and CVDs. GPA analysis revealed that all BCTs-CVDs trait pairs exhibited varying degrees of genetic overlap at a 5% FDR threshold, regardless of whether the genome-wide *r_g_* was significant (Supplementary Fig.2 and Supplementary Table 5). Notably, reticulocyte count (RET) and CAD showed a relatively high proportion of shared SNPs (proportion of association ratio (PAR) = 25.4%), consistent with their significant genetic correlation (*r_g_* = 0.107). Among the 138 BCTs-CVDs pairs with non-significant genome-wide *r_g_*, the overlap between CAD and MSCV was particularly pronounced, with a shared genetic overlap of 24.1%. As for IRF-VTE, as a significant genome-wide *r_g_* trait pair, the proportion of pleiotropic SNPs is only 19.0%, which is lower than MSCV-CAD. In summary, we confirmed the presence of shared genetic structure in all trait pairs using LAVA and GPA approaches. Although some trait pairs did not show evidence of genome-wide *r_g_*, possibly due to confounding effects, our analyses highlight potential genetic links that contribute to the complex relationship between BCTs and CVDs.

### Mendelian Randomization between BCTs and CVDs

To investigate vertical pleiotropy between 29 BCTs and 6 CVDs, we employed Latent Heritable Confounder Mendelian Randomization (LHC-MR) to evaluate causal relationships. The forward analysis identified seven causal relationships that achieved Bonferroni-corrected significance, including positive associations for RDW-VTE, basophil percentage of leukocytes (BASO_P)-AF, and BASO_P-CAD. For example, an elevated RDW_CV was associated with an increased risk of VTE (OR = 1.08), whereas a higher IRF was linked to a reduced risk of CAD (OR = 0.91), indicating negative causal relationships. The reverse analysis uncovered twelve positive causal relationships, such as AF, with an increased MCV (OR = 1.52). In contrast, four pairs showed negative causal relationships, notably VTE associated with a decreased RDW_CV (OR = 0.71). Remarkably, only the pair PCT-CAD exhibited a bidirectional causal relationship. These findings suggest that although some trait pairs demonstrate distinct causal connections, most associations between BCTs and CVDs are not driven by underlying causal factors(Fig. 2, Supplementary Fig. 3, Supplementary Table 6 and Supplementary Table 7).

**Figure 2:**
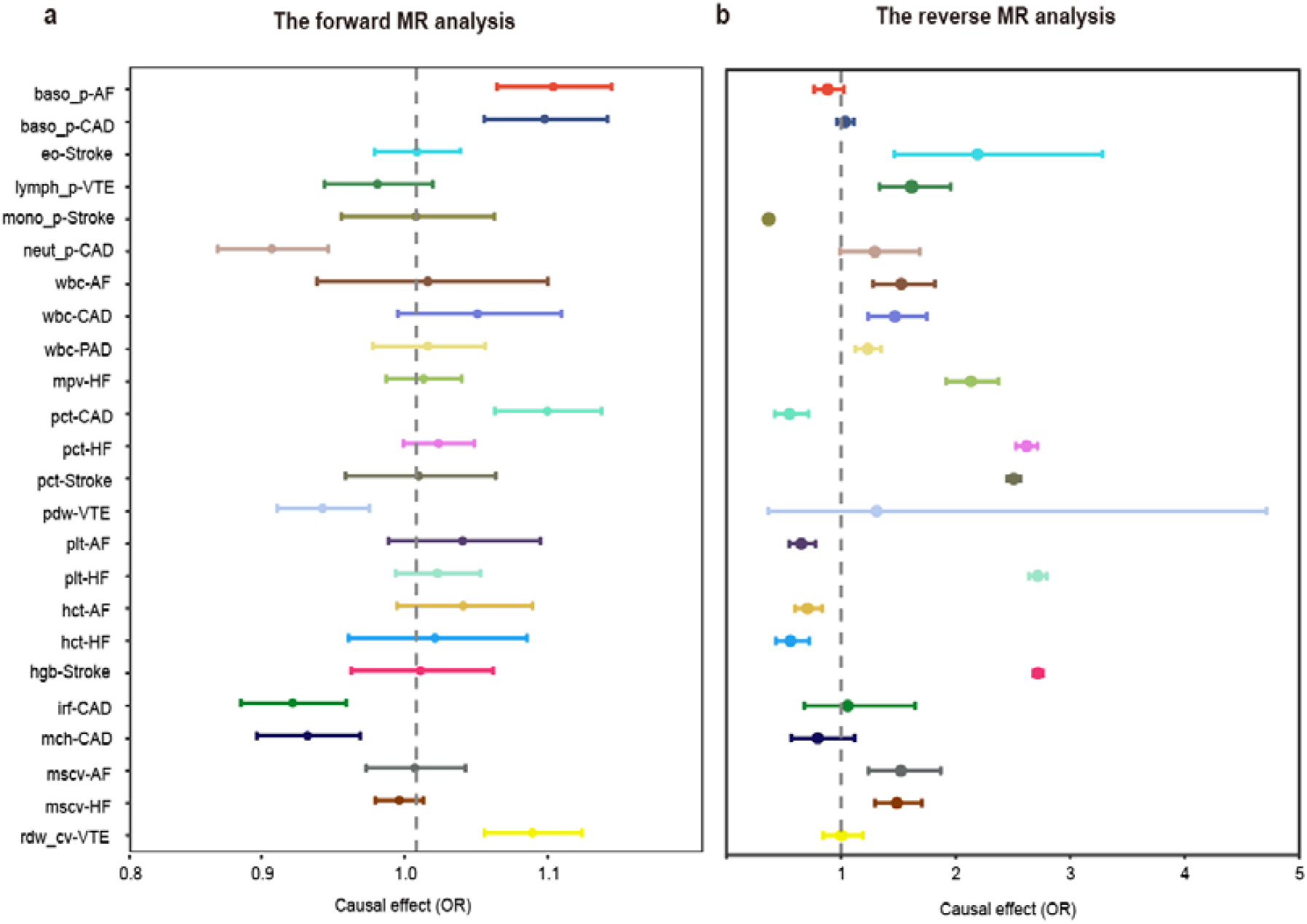
Forest plot of the bidirectional causal relationship between BCTs and CVDs. LHC-MR analysis was used to detect (a) forward (BCTs → CVDs) and (b) reverse (CVDs → BCTs) causal effects. Estimates and 95% confidence intervals are shown as dots and error bars, respectively.

### Shared Loci between BCTs and CVDs

Although the above three approaches have enhanced our understanding of the general pleiotropy between BCTs and CVDs, they still need to elaborate on the association patterns of individual loci and clarify their shared genetic architecture from the perspective of vertical and horizontal pleiotropy. To fill this gap, we employed the Pleiotropic Analysis under the Composite Null Hypothesis (PLACO) to identify horizontal pleiotropic SNPs mediating the association between BCTs and CVDs. A total of 571,782 SNPs (*P* < 5×10^-8^) were identified as significant pleiotropic variants across all BCT-CVD pairs (Fig. 3 and Supplementary Table 8). Notably, the PLT-CAD pair exhibited the most pleiotropic associations with 176 pleiotropic SNPs, followed closely by RBC-CAD (173 pleiotropic SNPs) and MSCV-CAD (168 pleiotropic SNPs), consistent with the significant genetic overlap between these trait pairs. Subsequently, 571,782 pleiotropic SNPs were clustered into 13,697 genomic loci in 580 unique chromosomal regions using functional mapping and annotation (FUMA). Specifically, 11,810 loci were associated with BCTs, 3,904 with CVDs, and 2,622 were shared between BCTs and CVDs. Notably, 12 chromosomal regions were shared by at least half of the trait pairs, including regions 9q34.2, 12q24.12, 12q24.31, and 19p13.2. We also observed mixed directions of allelic associations, with 6,721 top SNPs (49.1%) consistently associated with specific traits, indicating that these variants can either simultaneously reduce (3,185 SNPs) or increase (3,536 SNPs) the number/percentage of BCTs and CVDs risks. In contrast, the remaining 50.9% of the top SNPs showed opposite associations with specific traits, suggesting the presence of different biological mechanisms.

**Figure 3:**
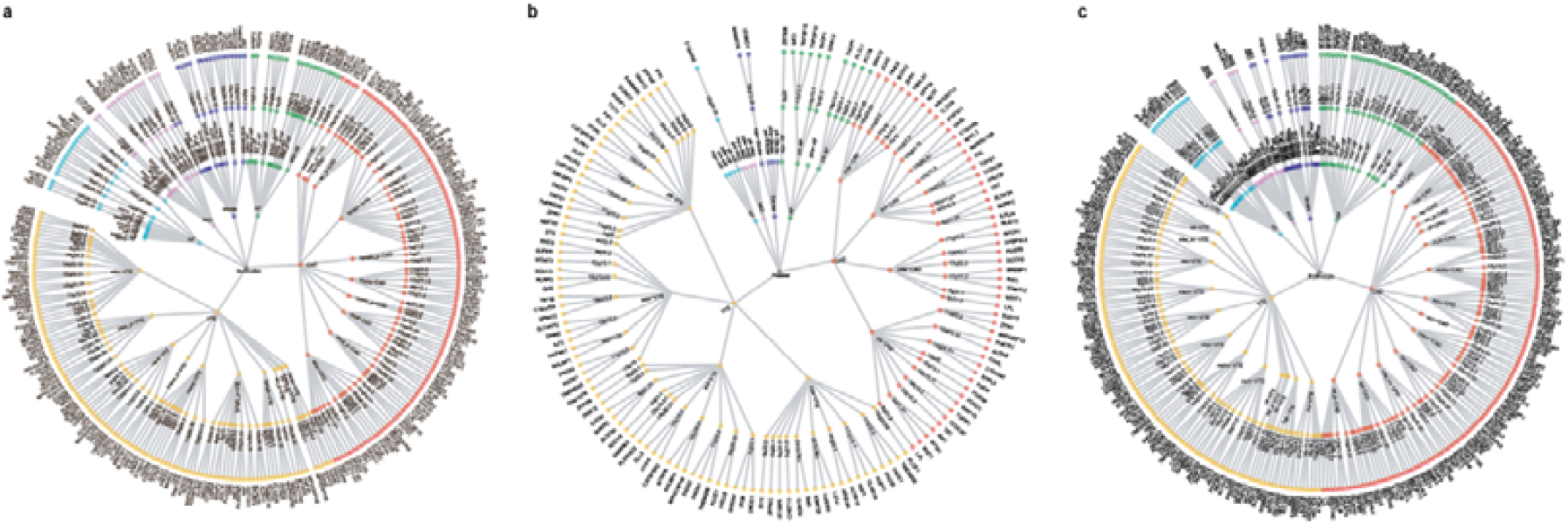
Overall landscape of the pleiotropic associations across three types of BCTs and six major CVDs. The three circular dendrograms included three BCTs (central points, including [a] leukocytes, [b] platelets, and [c] erythrocytes) and six CVDs (first circle), resulting in 66, 24, and 84 trait pairs (second circle). A total of 156, 81, and 192 pleiotropic loci were identified in 43, 13, and 58 trait pairs for (a) leukocyte-CVDs, (b) platelet-CVDs, and (c) erythrocyte-CVDs, respectively (third circle). Significant pleiotropic genes were further identified by MAGMA and EMAGMA, and a total of 936 pleiotropic genes co-localized for paired traits. For trait pairs with more than three pleiotropic genes, we only displayed the top three pleiotropic genes according to the priority of candidate pleiotropic genes (fourth circle).

For functional annotation, we utilized ANNOVAR and discovered that 8,201 (59.9%) of the variants were intronic, 3,749 (27.4%) were intergenic, and 523 (3.8%) were exonic. Among these, the index SNP rs1613662 at the 19q13.42 locus (*P_PLACO_* = 9.26 ×10^-10^ for basophil count BASO-VTE and PLT-VTE) was related to an adipose visceral omentum eQTL (*P_Adipose_Visceral_Omentum_* = 1.49×10^-5^), encoding the collagen receptor GPVI/FcRγ, primarily involving *GP6* variants (Supplementary Table 10). This receptor interaction with subendothelial collagen exposed after vessel wall injury can promote platelet activation and aggregation, thus affecting the risk of VTE^27^. Additionally, combined annotation-dependent depletion (CADD) scores revealed that 1,031 SNPs had scores greater than 12.37, with rs116843064 in the 19p13.2 region having the highest CADD score of 33. A total of 484 top SNPs were found to potentially affect transcription factor binding and correlate with gene target expression. Among these, six SNPs, including rs4987082, rs12440045, rs6511703, rs1214761, rs9810512, and rs6882088, had the highest credibility for regulatory functions (RegulomeDB score: 1a). We further performed colocalization analysis on 13,697 potential pleiotropic loci, of which 1,709 (12.5%) showed strong colocalization evidence (PPH4 > 0.7). Notably, the 12q24.31 locus affected 46 trait pairs, with PP.H4 values ranging from 0.740 to 0.995, demonstrating significant pleiotropy.

### Candidate pleiotropic genes by position mapping

We further integrated these pleiotropic SNPs to the gene level using multi-marker analysis of genomic annotation (MAGMA) and identified 10,106 significant pleiotropic genes (1,973 unique genes) (*P* < 1.63×10^-8^) (Fig. 3, Supplementary Table 12). Notably, 1,453 (73.6%) of these genes were widely shared between two or more trait pairs. For example, *ATXN2*, *ACAD10*, *ALDH2, CUX2, BRAP, SH2B3, and MAPKAPK5* were identified as significant pleiotropic genes in 69, 67, 64, 61, 58, 58 and 56 trait pairs, respectively. These genes are located in the 12q24.11-12q24.13 region, known for its pleiotropic effects on PCT and various cardiometabolic traits, including fasting glucose, blood pressure, and obesity-related traits (body mass index (BMI) and waist-to-hip ratio (WHR)). Depleting the cytoplasmic protein encoded by *ATXN2* has resulted in defective platelet aggregation and dysregulation of hemostatic processes^28^. Additionally, polymorphisms in *ATXN2* significantly affect the kynurenine level in erythrocytes^29^ and are critical loci for systolic (SBP) and diastolic blood pressure (DBP), as well as mean arterial pressure (MAP)^30^. Consequently, *ATXN2* plays a pivotal role in the onset of various CVD events by regulating blood pressure. To ensure the robustness of our findings, SNPs were positionally remapped using the SNP2GENE function of FUMA, which verified 88.5% of the genes identified by MAGMA (Supplementary Table 10). Further comparisons of the PLACO-based MAGMA results with single trait GWAS data for BCTs and CVDs revealed 855 (8.5%) novel genes for BCTs, 4,938 (48.9%) novel genes for CVDs, and 229 genes that overlapped between BCTs and CVDs (Supplementary Table 12).

### Candidate tissue-specific pleiotropic genes by eQTL mapping

To elucidate how the above SNPs affect gene expression in a tissue-specific manner, we first used LDSC-specific expression (LDSC-SEG) to determine the tissue association corresponding to each trait. We found that BCTs-related traits were mainly significantly enriched in Adipose_Visceral_(Omentum), Cells_EBV-transformed_lymphocytes, Lung, Small_Intestine_Terminal_Ileum and Whole_Blood tissues (FDR < 0.05) (Supplementary Fig. 4, Supplementary Table 15A). In CVDs, AF showed significant enrichment in heart-related tissues (Heart_Atrial_Appendage and Heart_Left_Ventricle), while CAD showed considerable enrichment in artery-related tissues (Artery_Aorta, Artery_Coronary, and Artery_Tibial). Subsequent LDSC-SEG chromatin analysis showed that BCTs were mostly significantly enriched in primary cells from peripheral blood (Supplementary Table 15B). In CVDs, AF was mainly enriched in the fetal heart, and CAD was significantly enriched in the aorta and coronary artery, confirming the multi-tissue gene expression results.

To overcome the limitation of the MAGMA approach (assigning SNPs to the nearest gene without considering functional association), we employed eQTL Multi-marker Analysis of GenoMic Annotation (eMAGMA) to link tissue-specific cis-eQTL information to genes to generate more biologically meaningful results (Supplementary Table 16). From an analysis of 10 selected tissues, we identified 2,328 unique pleiotropic genes achieving Bonferroni-corrected significance, each highly enriched in at least one specific tissue. Notably, 1,875 of these genes (80.5%) were present in two or more trait pairs. Among them, genes such as *ABO* (n=125), *TMEM116* (n=99), *ALDH2* (n=95), and *MAPKAPK5* (n=87) appeared in over half of the trait pairings, each demonstrating specific tissue enrichments. In particular, the *ABO* gene is predominantly found in liver and visceral adipose tissues; *TMEM116* in the artery tibial and visceral adipose tissues; *ALDH2* in both artery tibial and artery aorta tissues; and *MAPKAPK5* in adipose subcutaneous and visceral adipose tissues. Notably, the *ABO* gene affects the serum levels of von Willebrand factor (vWF) and soluble E-selectin^31^, which are involved in thrombosis and endothelial dysfunction, respectively, and are critical for the development of cerebrovascular injury and atherosclerosis^31,32,33,34^. We further used the single-trait GWAS results to conduct transcriptome-wide association study (TWAS) to identify tissue-specific pleiotropic genes for complex traits and found 5,609 novel genes for BCTs and 20 novel genes for CVDs (Supplementary Table 17). Notably, genes such as *ABO* and *ALDH2* were identified in more than half of the trait pairs, and both eQTL-based strategies highlighted that *ALDH2* in the 12q24.12 region was associated with a common molecular mechanism for BCTs and CVDs, which may mediate CVDs pathophysiology by affecting the metabolism of reactive aldehydes under oxidative stress. However, the exact role of *ALDH2* in the blood phenotype remains to be elucidated. In addition, we further validated 81.7% of the genes identified by eMAGMA using eQTL mapping using FUMA (Supplementary Table 10).

In our study, MAGMA and eMAGMA analyses identified 5,063 pleiotropic genes, of which 936 (313 unique) exhibited strong evidence of colocalization and were included in subsequent analyses (Supplementary Table 8 and Supplementary Table 12). Notably, 194 of these genes demonstrated pleiotropic effects across two or more BCT-CVD trait pairs, including significant genes such as *ALDH2*, *PHETA1*, and *MAPKAPK5* (all located at 12q24.12), *CCDC92* (12q24.31), and *ZNF664* (12q24.31). Interestingly, the degree of pleiotropy varied significantly among some genes across different categories of trait pairs. For instance, of the 18 trait pairs associated with *ACAD10*, 15 involved Leukocyte-CVD pairs, while the remaining 3 were Erythrocyte-CVD. Similarly, *CCDC92* appeared in 20 trait pairs, with 15 categorized as Erythrocyte-CVD, accounting for 17.9% of the total Erythrocyte-CVD. Conversely, *CCDC92* featured less frequently in Leukocyte-CVD (4 pairs, representing 6.1% of all such pairs) and Platelet-CVD (only 1 pair, or 4.2% of these pairings).

Therefore, considering that the association of genes with CVDs may be influenced by BCT type, we categorized all BCTs-CVDs trait pairs into three groups based on BCT types, including Leukocytes-CVDs, Platelets-CVDs, and Erythrocytes-CVDs, with pleiotropic gene counts of 174, 117, and 164, respectively. Among the Leukocytes-CVDs trait pairs, *ACAD10,* one of the specific genes, is not associated with Leukocytes-AF. *ACAD10* is critical in mitochondrial fatty acid β-oxidation, influencing leukocyte functions such as proliferation, cytokine production, and adhesion molecule production. It is also an essential energy source for cardiomyocytes, underscoring its significant role in CVD pathophysiology^35^. Among the Platelets-CVDs pairs, *TNFSF12* (*TWEAK*) was notably associated with PCT and AF. The interaction of *TNFSF12* with the fibroblast growth factor-inducible molecule 14 (Fn14) activates signaling pathways essential for vascular and cardiac remodeling, pivotal in acute and chronic CVDs^36^. Furthermore, studies have linked serum *TWEAK* levels to PLT, highlighting clinical relevance^33,37^. For the Erythrocytes-CVDs pairs, *CCDC92* was the most frequently pleiotropic gene, which affects the occurrence of cardiovascular events by regulating adipose tissue distribution and insulin sensitivity, highlighting its broad impact on metabolic and cardiovascular health^38,39^.

### Potential shared biological mechanism between BCTs and CVDs

We performed MAGMA gene set analysis to explore specific biological pathways or cellular functions implicated in the BCTs-CVDs pair’s genetic etiology. A total of 1,599 pathways were significantly enriched after Bonferroni correction (*P* < 3.00 × 10^-8^), including 169 Gene Ontology Biological Process (GO BP) terms, 30 KEGG pathways, and 33 Reactome pathways (Supplementary Table 18). Since most genes have obvious trait specificity and participate in different biological pathways, we still divide the pathways enriched by BCTs-CVDs into three categories to observe whether they have specific and reliable results. Of these, 992 were associated with the Leukocytes-CVDs pairs, and more than half of these involved pathways were related to the regulation of immune system processes, immune responses, and cellular activation, affecting all leukocyte phenotypes except basophil-related parameters. In contrast, the Platelets-CVDs pairs were enriched for only 77 pathways, mainly related to hemostasis and platelet activation, signaling, and aggregation. VTE showed strong correlations with all four platelet parameters in these specific pathways. For the Erythrocytes-CVDs pairs, 530 pathways were enriched, mainly involving vascular and circulatory system development, with AF and CAD being more highly associated with these terms than other CVDs.

Further enrichment analysis of overlapping genes in MAGMA and eMAGMA analyses using Metascape confirmed that the Leukocytes-CVDs related gene set was mainly involved in the inflammatory response, the Platelets-CVDs related gene set focused on platelet activation and aggregation, and the Erythrocytes-CVDs related gene set was mainly engaged in the regulation of hematopoietic progenitor differentiation, which is highly consistent with the results of the MAGMA gene set analysis(Supplementary Table 19). These findings revealed the common molecular genetic mechanisms behind the extensive multi-gene overlap between the three types of BCTs and CVDs, highlighting important pathways that may become therapeutic targets for treating these diseases.

### Identification of pathogenic proteins and drugs in cross-sectional traits

To investigate the associations between plasma protein expression and disease risk, we employed summary-data-based Mendelian Randomization (SMR). We sourced blood cis-pQTL data from the UK Biobank Pharmaceutical Proteomics Project (UKB-PPP) and the deCODE Health study. After excluding entries that failed the heterogeneity in dependent instruments (HEIDI) test, underwent multiple SNP-SMR sensitivity analyses, and surpassed the Bonferroni correction threshold, we identified 1,589 causal proteins, of which 1,534 were associated with BCTs and 55 with CVDs. Among these, 49, 62, and 101 overlapping proteins were associated with leukocytes, platelets, and erythrocytes, respectively, from deCODE Health and UKB-PPP studies, with an additional eight proteins linked to CVDs. Remarkably, our analysis uncovered only three pleiotropic proteins across seven distinct trait pairs, specifically SERPINE2 and TNFSF12 in Platelets-CVDs pairs and GP6 in both Platelets-CVDs and Erythrocytes-CVDs pairs. Subsequent colocalization analysis confirmed that the GP6 gene polymorphic variant rs892090 demonstrated strong colocalization in all pertinent trait pairs except MPV-VTE (Supplementary Table 20).

## Discussion

We discovered significant genetic overlaps between BCTs and CVDs through comprehensive pleiotropic analyses of large-scale GWAS summary statistics. However, evidence for a direct causal relationship remains limited. Our studies identified 571,782 pleiotropic variants and 313 unique pleiotropic genes, which show varied CVD risks across different BCT subgroups, including leukocytes, platelets, and erythrocytes. We further explored the distinct characteristics of these blood cell phenotypes, revealing that specific pathways play key roles in their pathophysiology. For platelet-CVD subgroups, platelet activation is central; for erythrocyte-CVD subgroups, the regulation of hematopoietic progenitor differentiation is crucial; and for leukocyte-CVD subgroups, the inflammatory response is pivotal. Additionally, our research has identified GP6, TNFSF12, and SERPINE2 as potential therapeutic targets for these conditions. These findings underscore the value of BCT parameters as biomarkers for assessing CVD risk and provide valuable insights into the genetic underpinnings of these associations.

We identified a substantial genetic overlap between BCTs and CVDs using diverse analytical methodologies, including LDSC, GPA, and LAVA. Our analysis indicated significant genome-wide genetic correlations for 8.6% of trait pairs, corroborating previous phenotypic associations and confirming a shared genetic foundation. Although genome-wide *r_g_* (averaging shared SNP effects across the genome) may obscure specific genetic associations within genomic regions, we utilized LAVA to pinpoint multiple specific genomic areas. We observed local-*r_g_s* for all trait pairs within these regions, indicating confounding effects that are not evident in genome-wide estimates. For instance, despite a genome-wide *r_g_* near zero according to LDSC, both BASO_P-AF and BASO-CAD demonstrated approximately equal numbers of positively and negatively correlated genetic regions. This nuance underscores significant genetic correlations that broader statistical frameworks might overlook. Our GPA analysis further confirmed substantial polygenic overlap among all trait pairs, highlighting the complex genetic interplay between BCTs and CVDs. In conclusion, our study reveals an extensive shared genetic liability between BCTs and CVDs, suggesting that their relationships are more robust and intricate than previously recognized.

The LHC-MR method explored the causal relationship at the level of vertical pleiotropy, which can reveal part of the common genetic mechanisms between BCTs and CVDs. We identified 23 putative causal relationships, especially positive ones in RDW_CV-VTE, Stroke-eosinophil count (EO), and Stroke-PCT, consistent with previous studies^20,40^. The causal link for RDW_CV-VTE may be the reduced deformability and increased aggregation of red blood cells, elevating viscosity and impeding blood flow, thereby increasing VTE risk. Conversely, our findings did not confirm the previously reported associations between high neut and increased risks of AF, HF, PAD, and Stroke, as suggested by Jiao *et al*^41^. This discrepancy could stem from the significant sample overlap between BCTs and CVDs GWAS samples in the UK Biobank, a typical challenge in two-sample Mendelian randomization analyses. Our LHC-MR approach addressed and mitigated genetic confounding due to this sample overlap and clarified the exposure-outcome relationships. Additionally, we report for the first time a positive association between HF and several BCTs, such as MPV, MSCV, PCT, and PLT, and a negative association with HCT. However, the negative relationship between AF and PLT is consistent with previous studies. Specifically, a meta-analysis by Alexander *et al*. indicated a significant reduction in PLT in AF patients, potentially influenced by factors such as AF type, smoking status, and geographic region^42^. We found no causal associations between the remaining trait pairs, and the limited causal evidence suggests that the BCTs-CVDs trait pair associations are less driven by causality and may be primarily mediated by horizontal pleiotropy.

Although we discovered that BCTs and CVDs share a common genetic foundation, it remains unclear whether the association between them occurs via horizontal or vertical pleiotropy. We first investigated the horizontal pleiotropy level and identified multiple pleiotropic loci associated with BCTs and CVDs, among which important loci such as 9q34.2, 12q24.31, and 12q24.12 affected more than 50% of the trait pairs. Recent research has underscored the pleiotropic effects of genes within the 12q24.12 region on CVDs^43^. Notably, the gene *ALDH2*, encoding aldehyde dehydrogenase 2, is associated with CAD, blood pressure regulation, and alcohol-induced cardiac dysfunction^44^. This enzyme is essential for metabolizing both endogenous and exogenous aldehydes; its dysfunction leads to aldehyde accumulation, markedly increasing the risk of specific CVDs. Furthermore, *ALDH2* modulates atherosclerosis-related risk factors such as foam cell formation and macrophage effervescence through non-enzymatic pathways^45^. Another critical gene in this region, *SH2B3* (also known as lymphocyte adaptor protein or *LNK*), regulates cytokine signaling and cell proliferation^46^. In mouse models, targeted deletion of *LNK* disrupts the negative feedback regulation of various pathways, affecting hematopoiesis and enhancing thrombopoietin (TPO) signaling^47^. This disruption leads to increased thrombosis and atherosclerosis, thereby elevating the incidence of CVD events. Consistently, studies have linked the *LNK* rs3184504 (T allele, *R262W*) variant with an increased risk of CAD and thrombotic stroke^48^. HyPrColoc results confirm that rs3184504 is a common pathogenic variant across CAD, VTE, and 19 other BCTs, highlighting the pleiotropic role of *SH2B3*. The gene *MAPKAPK5*, which encodes the MK5 protein—a serine/threonine kinase—also shows high expression levels in the human heart^49^. MK5 plays a role in endothelial cell migration and angiogenesis^50^ and influences erythrocytes’ structure, metabolism, and ion channels^51^, thus affecting susceptibility to atherosclerosis^52^. In conclusion, the identified pleiotropic loci associated with BCTs and CVDs reveal a strong genetic interrelation between these traits, providing deeper insights into their complex genetic associations.

In our study, we employed two gene mapping strategies to associate candidate SNPs with genes that influence both BCTs and CVDs. Specifically, among the Leukocytes-CVDs trait pairs, *ALDH2*, *MAPKAPK5*, and *ACAD10*, all located at 12q24.12, were identified as significant pleiotropic genes. *ACAD10*, in particular, plays a crucial role in activating mitochondrial fatty acid oxidation (FAO)^53^, which is pivotal in developing atherosclerosis through *NLRP3* inflammasome activation and IL-4-induced macrophage polarization^54^. Notably, the FAO inhibitor trimetazidine effectively prevents foam cell formation from macrophages originating from circulating monocytes in the arterial intima^55^. About Platelets-CVDs trait pairs, *TNFSF12* (17p13.1) has emerged as key pleiotropic genes. SMR studies have demonstrated a significant association between *TNFSF12* protein levels and PCT-AF. *TNFSF12*, also known as *TWEAK*, activates the JAK2/STAT3 pathway via the Fn14 receptor, inducing hypertrophy in HL-1 atrial myocytes—a vital adaptive change in AF development^56^. This pathway also influences PLT and vascular permeability^33^. Given their potential, the drugs BIIB-023 and RO-5458640, currently indicated for lupus nephritis and rheumatoid arthritis, respectively, warrant further investigation in clinical trials to evaluate their efficacy in treating PCT and AF. Regarding Erythrocytes-CVDs trait pairs, *ZNF664* and *CCDC92*, both located in the 12q24.31 region, are critical. *CCDC92*, a coiled-coil domain protein, is intimately associated with lipid metabolism and insulin sensitivity^38^. Insulin resistance, influenced by *CCDC92*, leads to coronary endothelial dysfunction and promotes vascular smooth muscle cell proliferation via insulin-like growth factor receptors, exacerbating atherosclerosis^57^. At the same time, no studies have directly linked *CCDC92* with erythroid characteristics; *ZNF664* and *CCDC92* exhibit co-regulated expression patterns and engage in nearly identical molecular pathways^58,59^.

Shared genetic determinants indicate common biological pathways underlying Leukocytes-CVDs trait pairs. Our pathway analysis revealed that inflammatory responses are critical, with *TNFSF12* playing a significant role. Specifically, inflammatory reactions within the vessel wall, coupled with oxidative stress from endothelial dysfunction, are pivotal in the progression of atherosclerosis^60^. For example, circulating monocytes infiltrate fatty streaks and differentiate into macrophages with pro-inflammatory properties^61^. These macrophages then recruit inflammatory factors, amplifying local inflammatory responses. A notable mediator, Irg1, exacerbates inflammation by promoting the formation of neutrophil extracellular traps (NETs) and activating the *NLRP3* inflammasome in macrophages, enhancing IL-1β release ^62^. In analyzing Platelets-CVDs trait pairs, we focused on key processes such as hemostasis, platelets activation, and aggregation involving *GP6*. Platelets, derived from megakaryocytes, are fundamental to the pathogenesis of coronary thrombosis and atherosclerosis^63^. Peptide hormone receptors on platelets may trigger thrombosis^64^, while the chemokines they secrete play roles in both inflammation and hemostasis^65^. Additionally, oxidative stress and the production of reactive oxygen species (ROS) activate platelets, profoundly affecting CVD pathogenesis, especially in older individuals. Regarding Erythrocytes-CVDs trait pairs, pathways involved in vascular development and the circulatory system are crucial, with *TNFSF12* also implicated. Erythrocytes primarily influence blood viscosity, impacting the friction exerted on the arterial wall, a key factor in maintaining systemic arterial pressure post-birth and promoting vasoconstriction^5^. Pathologically, collisions of erythrocytes with the arterial wall can lead to local retention and hemolysis of their membrane lipids, which are associated with both the early and late stages of atherosclerosis^66,67^. From a classification perspective, our study elucidates how different pathways mediate the association between BCTs and CVDs, highlighting that targeted regulation of key genes could significantly reduce disease risk.

Using SMR and colocalization analysis, we identified three significantly shared proteins—GP6, TNFSF12, and SERPINE2—across seven trait pairs. Both GP6 and TNFSF12 are key target proteins currently under clinical investigation. GP6 encodes platelets glycoprotein VI, essential for collagen-dependent activation, signal transduction, and full platelets activation, adhesion, and aggregation^68^. Variants in GP6, such as the common variant rs1613662, are associated with increased VTE risk and alterations in platelets function, impacting CVDs susceptibility. Although no studies have reported how GP6 participates in erythrocyte physiological processes, our analysis highlights that GP6 upregulation correlates with heightened VTE risk and elevations in 2 platelet parameters (MPV and PDW) and 4 erythrocyte parameters (HLR_P, HLR, RET_P, and RET). This suggests that the GP6 inhibitor, glenisomab, may represent a promising drug repurposing opportunity to mitigate excessive VTE risk and elevated platelet and erythrocyte parameters. Restingly, common CVD preventive drugs such as aspirin do not target the pathogenic proteins identified in our SMR analysis. This may be due to the inability of blood-based cis-eQTL analysis to capture anticoagulant factors produced by the liver. Moreover, the TNFSF12 inhibitor BIIB-023 could be repurposed to reduce the risk of PCT and AF complications. However, the underlying mechanisms remain under-explored, highlighting our proposal as novel and underscoring the need for further research.

In conclusion, this is the first study to characterize the pleiotropic effects of a range of BCTs and CVDs on a large scale and provide important insights into the genetic architecture widely distributed throughout the genome and the mechanisms behind common genetic diseases. These results have successively provided new clues at the level of pleiotropic SNPs and genes, biological pathways, potential drug proteins, and causal relationships and will facilitate further laboratory investigations and clinical studies.

## Limitations of the study

Our study presents several limitations. First, the GWAS datasets employed to investigate BCTs and CVDs may include cases with concurrent blood cell abnormalities and CVDs, potentially introducing bias into our analysis of genetic overlap. Second, the lack of rare variants in most GWAS datasets restricts our ability to explore pleiotropy between rare loci associated with BCTs and CVDs, which could deepen our understanding of the disease mechanisms. Third, while minimizing demographic bias, our focus on European populations limits our findings’ generalizability to other ethnic groups. Finally, the new pathway signals and gene loci identified require further validation through clinical cohort studies or animal models to bolster the robustness and credibility of our results.

## STAR ★ Methods

### Key resources table

| REAGENT or RESOURCE | IDENTIFIER |
| --- | --- |
| Deposited data |  |
| Blood cell traits | [ftp://ftp.sanger.ac.uk/pub/project/humgen/summary_statistics/UKB<br>B_blood_cell_traits/](ftp://ftp.sanger.ac.uk/pub/project/humgen/summary_statistics/UKB_B_blood_cell_traits/) |
| Atrial fibrillation | https://www.ebi.ac.uk/gwas/studies/GCST006414 |
| Coronary artery disease | https://cvd.hugeamp.org/datasets.html |
| Venous thromboembolism | https://www.decode.com/summarydata/ |
| Heart failure | https://www.ebi.ac.uk/gwas/studies/GCST009541 |
| Peripheral artery disease | https://cvd.hugeamp.org/datasets.html |
| Stroke | https://www.ebi.ac.uk/gwas/studies/GCST90104539 |
| Software and algorithms |  |
| LDSC (v1.0.1) | https://github.com/bulik/ldsc |
| MiXeR (v1.3) | https://github.com/precimed/mixer |
| LAVA (v0.1.0) | https://github.com/josefin-werme/LAVA |
| LHC-MR (v0.0.0.9000) | https://github.com/LizaDarrous/lhcMR |
| PLACO (v0.1.1) | https://github.com/RayDebashree/PLACO |
| FUMA (v1.5.4) | http://fuma.ctglab.nl/ |
| HyPrColoc(v1.0) | https://github.com/jrs95/hyprcoloc |
| MAGMA (v.1.08) | https://ctg.cncr.nl/software/magma |
| e-MAGMA | https://github.com/eskederks/eMAGMA-tutorial |
| TWAS | http://gusevlab.org/projects/fusion/ |
| SMR (v1.31) | https://yanglab.westlake.edu.cn/software/smr/ |
| COLOC (v5.2.1) | https://github.com/chr1swallace/coloc |
| R (v.4.1.3) | https://www.r-project.org/ |

### Resource availability

#### Lead contact

Further information and requests for resources and reagents should be directed to and will be fulfilled by the lead contact, Luke Kong.

### Materials availability

This study did not generate new unique reagents.

### Data and code availability

The study used only openly available GWAS summary statistics on blood cell traits and cardiovascular diseases that have originally been conducted using human data. GWAS summary statistics on BCTs are available at ftp://ftp.sanger.ac.uk/pub/project/humgen/summary_statistics/UKBB_blood_cell_traits/. GWAS summary statistics on AF, HF, and Stroke are available at the GWAS Catalog (GCST90104539, GCST009541, and GCST90104539). GWAS summary statistics on CAD and PAD are publicly available for download at the Cardiovascular Disease Knowledge Portal (CVDKP) website: https://cvd.hugeamp.org/datasets.html. GWAS summary statistics on VTE are obtained from the deCODE genetics website: https://www.decode.com/summarydata/. All data are publicly available and listed in the key resources table. No unique datasets or code were generated for this study. Any additional information required for reanalysis of the data reported in this article is available from the primary contact on request.

### Method details

#### Study Design

Figure 1 presents the workflow for this study

#### Data selection and quality control

Summary statistics for 29 BCTs were retrieved from a genome-wide association study of 408,112 participants of European ancestry in the UK Biobank cohort, which included data on leukocytes, platelets, and erythrocytes^16^. Additionally, we integrated GWAS summary data on six major CVDs for individuals of European ancestry with sample sizes exceeding 50,000 to ensure robust statistical power. Specifically, summary data for AF were derived from a large-scale meta-analysis including six contributing studies: the Nord-Trøndelag Health Study (HUNT), deCODE, the Michigan Genomics Initiative (MGI), DiscovEHR, UK Biobank, and the AFGen Consortium, encompassed 60,620 cases and 970,216 controls of European ancestry^69^. Summary data for CAD were derived from GWAS summary statistics from a genome-wide meta-analysis by the CARDIoGRAMplusC4D Consortium and UK Biobank, including 181,522 cases and 984,168 controls of European ancestry^70^. Summary data for VTE were sourced from a genome-wide association study by Jonas Chouse et al., featuring 81,190 cases and 1,419,671 controls of European ancestry^71^. Summary statistics for HF were extracted from a GWAS meta-analysis by Sonia Shah et al., including data from 47,309 cases and 930,014 controls of European ancestry across 26 studies from the Heart Failure Molecular Epidemiology for Therapeutic Targets (HERMES) Consortium^72^. Summary statistics for PAD were obtained from a large-scale GWAS meta-analysis comprising 12,086 cases and 499,548 controls of European ancestry^73^. Summary statistics for stroke, involving 73,652 cases and 1,234,808 controls of European ancestry, were retrieved from the GlGASTROKE consortium^74^. Detailed information on these GWAS summary statistics and their original publication sources is available in Table 1.

We implemented stringent quality control measures to ensure the integrity of our GWAS summary data and facilitate a valid comparison between BCTs and CVDs. These included (i) aligning the data with the hg19 genome build based on the 1000 Genomes Project v3 Europeans reference; (ii) filtering out SNPs that either lacked a reference SNP ID (rsID) or had duplicated rsIDs; (iii) excluding non-biallelic SNPs, which do not have precisely two allele forms; (iv) removing SNPs with a minor allele frequency (MAF) below 0.01, ensuring that only commonly occurring variants are analyzed. Following these measures, we included only common SNPs in the analysis, totaling 6,923,169, available across all traits.

### Statistical Analysis

#### Genome-wide genetic correlations between BCTs and CVDs

We used LDSC to analyze the *h^2^_SNP_* and *r_g_* between 29 BCTs and 6 major CVDs. LDSC helps quantify the contributions of polygenic effects by examining the relationship between linkage disequilibrium scores from GWAS summary results and SNP test statistics^75^. Initially, univariate LDSC was used to assess the SNP-based heritability for each of the 29 BCTs and 6 CVDs, which measures the proportion of phenotypic variance explained by common genetic variants. For this analysis, we calculated LD scores using the 1000 Genomes Phase 3 European reference^76^, excluding variants within the major histocompatibility complex (MHC) region (CHR 6: 25–35 Mb) due to its complex LD structure. We then conducted bivariate LDSC to evaluate the genetic correlations among 174 BCTs and CVDs pairwise combinations. This step calculates the proportion of SNP-based heritability shared between two traits, normalized by the geometric mean of their heritability estimates, with correlation values ranging from -1 to +1 to indicate the direction of genetic effects. To adjust for multiple comparisons, we employed the FDR method, setting an FDR threshold of <0.05 to determine statistical significance.

We employed LDSC-SEG to explore whether the SNP heritability of 29 BCTs and 6 major CVDs is significantly correlated with tissue-specific gene expression, thereby identifying relevant tissues^77^. Utilizing multi-tissue gene expression data from the Genotype-Tissue Expression (GTEx) project and additional data from the Franke laboratory, we analyzed patterns across 53 tissues and 152 cell types^78^. Furthermore, we enhanced our validation process by incorporating chromatin-based annotations linked to six epigenetic marks: DNase hypersensitivity, H3K27ac, H3K4me1, H3K4me3, H3K9ac, and H3K36me3. These annotations included 93 tags from the Encyclopedia of DNA Elements (ENCODE) project and 396 tags from the Roadmap Epigenomics database, which validated our analysis. To determine the statistical significance of the coefficients for the identified tissues, we applied a FDR threshold of 0.05.

### Local genetic correlation analyses between BCTs and CVDs

To investigate local genetic correlations between 29 BCTs and 6 major CVDs within specific genomic regions, we utilized the LAVA. LAVA computed local-*r_g_s* for 2,495 semi-independent genetic loci, each approximately 1 Mb in size, identified by dividing the genome into blocks with minimal LD between them. The LD reference panel, derived from the 1000 Genomes Project Phase 3 of European ancestry^76^, excluded the MHC region (chr6:25-35 Mb). Initial univariate tests ascertained the local heritability of each trait using a stringent p-value threshold of < 1×10^-4^ to exclude loci with insignificant genetic correlation or univariate heritability. Subsequently, we performed 80,126 bivariate tests on loci and traits that exhibited significant univariate genetic signals. To address the conservative nature of the Bonferroni correction, we applied a FDR of 0.05 to identify significant associations.

For regions identified by LAVA with evidence of shared risk loci across multiple phenotypes, we conducted a multi-trait co-localization analysis using HyPrColoc^79^. This method builds on the COLOC framework and simultaneously assesses co-localization among several traits by calculating the PP of multiple traits co-localizing due to a single causal variant. A SNP with a PP exceeding 0.7 indicates a significant co-localization signal, pointing to shared causal mechanisms within the region.

### Genetic overlap analysis between BCTs and CVDs

To further explore the genetic overlap between BCTs and six major CVDs, we applied the GPA^80^. This method leverages GWAS summary results, using the intersection of P-values for each phenotype as input, and fits a GPA model based on the resulting P-value matrix data. GPA categorizes SNPs into four groups according to their association with the traits: SNPs unrelated to any traits (π00), related only to the first trait (π10), related only to the second trait (π01), and associated with both traits (π11). This classification helps estimate the proportions of SNPs and elucidates the causal effects on the phenotypes. The statistical significance of the genetic overlap is assessed using a Likelihood Ratio Test (LRT), which tests the fit of the four-group model against a model of independent effects. GPA assumes P-values from null SNPs follow a uniform distribution, while those from non-null SNPs follow a Beta distribution. The PAR, defined as π11/(π01 + π10 + π11), represents the proportion of SNPs associated with both traits relative to those associated with at least one trait^81^. To minimize the influence of LD on our GPA estimates, we performed LD pruning using PLINK^82^ with genotype data from the 1000 Genomes Project Phase 3 for European ancestry, selecting relatively independent SNPs^76^. We then applied the FDR method to correct for multiple testing, with an FDR threshold of <0.05 to determine statistical significance.

### Mendelian randomization analysis using LHCMR

We employed the LHC-MR method to explore potential causal relationships between 29 BCTs and 6 major CVDs^83^. Unlike traditional polygenic MR approaches that rely only on genome-wide significant SNPs, LHC-MR exploits all genome-wide variation for causal estimation, using structural equation models to relate genome-wide associations to traits and confounders. LHC-MR enhances the ability to estimate bidirectional causal effects, direct heritability, and confounding effects, as well as accommodate sample overlap. Moreover, we also applied traditional bidirectional MR models for sensitivity analysis, including MR Egger, weighted median, inverse variance weighted (IVW), simple mode, and weighted mode^84^. Estimates from LHC-MR are reported as odds ratios (ORs) with corresponding 95% confidence intervals (CIs). We applied a Bonferroni correction to rigorously evaluate causal relationships between trait pairs, setting a significance threshold at *P* < 1.44×10^-4^ (0.05 divided by 174 pairs, then divided by 2 to account for testing in both directions). A unidirectional causal relationship was established when the P-value in one direction was below the threshold, while in the opposite direction, it exceeded 0.05. Conversely, a bidirectional relationship was confirmed when the P-values in both directions were below 1.44×10^-4^.

### SNP-level Analysis

#### Pleiotropic SNPs Identified by PLACO

To assess the evidence for horizontal pleiotropy between 29 BCTs and 6 major CVDs, we utilized the PLACO. PLACO is a sophisticated statistical method designed to detect pleiotropic associations of each variant by evaluating the hypothesis that a SNP is associated with neither or just one of the traits^85^. This hypothesis is subdivided into four specific scenarios: (1) H00, indicating no association with either trait; (2) H10, suggesting an association exclusively with the first trait; (3) H01, indicating an association solely with the second trait; and (4) H11, representing a pleiotropic association with both traits. The test statistic is calculated as the product of the Z statistics for the SNPs for each trait, which is assumed to follow a mixture distribution. To ensure accuracy, SNPs with squared Z statistics (Z^2^) greater than 100 are excluded from analysis. SNPs are declared significantly pleiotropic at the genome-wide level if their *P*-values are less than 5×10^-8^.

#### Genomic loci definition and functional analysis

We employed the FUMA platform^86^ to identify and functionally annotate independent genomic loci and pleiotropic SNPs identified through PLACO. FUMA processes GWAS summary statistics to annotate and prioritize SNPs and genes, enhancing data interpretation through interactive visualizations^76^. Using European population data from the 1000 Genomes Phase 3 as a reference, we identified independently significant SNPs (*P* < 5 × 10^-8^, r^2^ < 0.6) from GWAS results. LeadSNPs were then defined from these SNPs based on mutual independence (r^2^ < 0.1), and genomic risk sites were determined where SNPs were in LD (r^2^ > 0.6). LD blocks within lead SNPs, separated by less than 500 kb, were combined into single sites, with the top lead SNP determined by the lowest P value in each site. We then evaluated the directional effects of these top SNPs by comparing Z-scores, considering them novel if inconsistent with any previously reported loci for BCTs or CVDs in original GWAS. For functional and pathogenic prediction of variants, we utilized ANNOVAR, applying metrics like the CADD score to predict the deleteriousness of variants based on 67 functional annotations. SNPs with a CADD score over 12.37 were considered harmful^87^. The regulatory potential of SNPs was assessed using RegulomeDB scores, ranging from 1a (strong evidence of regulatory function) to 7, with lower scores indicating higher activity^88^. Chromatin states, predicted by ChromHMM using five chromatin markers across 127 epigenomes, helped highlight the regulatory landscape of genomic regions^89^. Lastly, SNP-gene associations were analyzed through positional mapping within a 10 kb window of genes and eQTL mapping from GTEx v8 data to identify significant cis-SNP gene pairs within 1 Mb relevant to the studied traits.

#### Bayesian colocalization analysis

Bayesian colocalization analysis used the ’coloc’ R package on pleiotropic loci annotated by FUMA to identify shared causal variants within each locus^90^. This analysis assesses whether specific loci contain a causal variation that influences two traits simultaneously. We tested five hypotheses using COLOC to evaluate different scenarios of genetic influence at these loci^91^: PP0: no association with either trait; PP1: a causal variant affects only the first trait; PP2: a causal variant affects only the second trait; PP3: different causal variants affect each trait.; PP4: a common causal variant affects both traits. Loci with a posterior probability for hypothesis four (PP.H4) greater than 0.7 were deemed to exhibit significant colocalization. Within these loci, the SNP showing the highest PP.H4 was identified as the putative causal variant, suggesting a strong genetic link between the traits at this genomic location.

### Gene-level Analysis

#### Gene-based association analysis using MAGMA

To identify candidate pleiotropic genes based on the pleiotropic SNPs identified by PLACO and single-trait GWAS results, we employed the MAGMA. This analysis utilizes multiple linear principal component regression to map SNP annotations from GWAS data onto the genome, detecting gene-disease associations. MAGMA’s strength lies in aggregating evidence from multiple genetic variants within the same gene to enhance the detection of novel genetic signals. For analytical robustness, we restricted our analysis to protein-coding genes containing at least ten SNPs. Gene annotations were aligned with the Genome Reference Consortium Human Build 37 (hg19), and the analysis incorporated the 1000 Genomes Phase 3 European LD score reference panel^76^, focusing on 17,636 autosomal protein-coding genes. SNPs were systematically assigned to genes based on their location within the gene body or within a ±10 kb window surrounding the gene. Additionally, regions within the MHC (chr6:25-35 Mb) were excluded due to their complex linkage disequilibrium patterns. We applied a rigorous Bonferroni correction for multiple testing across 17,636 protein-coding genes and 174 trait pairs, setting a stringent significance threshold for the MAGMA analysis at P < 0.05/(17,636×174).

#### Investigation of the tissue-specific genes using EMAGMA and TWAS

To overcome limitations in identifying causal genes using traditional MAGMA, which assigns SNPs to genes within a genomic window, we implemented E-MAGMA^92^ for transcriptome-wide association analysis. This enhanced approach uses tissue-specific cis-eQTL information from the GTEx v8 to link SNPs more accurately with their putative genes, thereby improving the biological interpretability of gene-based association analyses in BCTs and CVDs. E-MAGMA, adhering to the statistical framework of MAGMA, utilizes multiple linear principal component regression models. We focused our analysis on ten GTEx v8 tissues previously identified as relevant to BCTs and CVDs, including arterial, adipose, and cardiac tissues. To mitigate analytical complexities, regions like the MHC (chr6:25-35 Mb) were excluded, and we employed Bonferroni-corrected significance thresholds tailored per tissue and trait pair, ensuring robust statistical validation.

Complementing our E-MAGMA analysis, we conducted TWAS^93^ using the Functional Summary-based Imputation (FUSION) approach. TWAS integrates gene expression data with single-trait GWAS results to uncover tissue-specific genetic influences. We employed various prediction models, including Best Linear Unbiased Prediction (BLUP), Least Absolute Shrinkage and Selection Operator (LASSO), Elastic Net, and Bayesian Sparse Linear Mixed Model (BSLMM), selecting the model that provided the best prediction accuracy for each tissue. Adjustments for multiple comparisons were made using the Bonferroni method across each tissue type to ensure the statistical rigor of our findings.

#### Pathway-level analysis using MAGMA and Metascape

We used MAGMA gene set analysis to elucidate the biological relevance of pleiotropic genes identified from overlaps detected by MAGMA and subsequent pathway enrichment annotation^94^. This involved a competitive gene set enrichment analysis using MAGMA, which assesses whether genes within specific sets exhibit stronger associations with the phenotype than those in other gene sets across the genome. Our analysis included 9,398 gene sets from the Molecular Signatures Database (MSigDB, version 2023.1), particularly focusing on the C2: Reactome Pathways and C5: GO Biological Processes and various biological process sets. We applied a stringent Bonferroni correction for multiple testing, setting the threshold at *P* = 0.05 / (7744 + 186 + 1654) / 174 = 3.00×10^-8^ to ensure robust statistical validity.

In addition, we used Metascape to perform pathway enrichment analysis on genes significantly identified in MAGMA and E-MAGMA analyses to elucidate the biological functions and pathways of pleiotropic genes associated with BCTs and CVDs. Metascape^95^ integrates functional enrichment analysis, interaction analysis, gene annotation, and member search functions to facilitate comprehensive bioinformatics analysis of bulk genes. For pathway enrichment analysis, we used the following ontology sources: GO annotation, KEGG, and Reactome pathways. P values below 0.01 were considered statistically significant.

#### Proteome-wide Mendeian Randomization analysis using SMR and Colocalization analysis

To explore potential common pathogenic factors at the proteomic level between 29 BCTs and 6 major CVDs, we employed SMR^96^. We analyzed disease GWAS data and pQTL data from the deCODE Health study and the UKB-PPP. The deCODE study measured 4,719 plasma proteins in 35,559 Icelandic individuals using the SomaScan platform, while the UKB-PPP assessed 2,940 proteins in 34,557 European individuals using the Olink Explore platform. Instrumental variables for SMR were selected based on genome-wide significant SNPs (P < 5 × 10^-8^) located within ±1 Mb of the transcription initiation sites of target genes. SMR is a novel MR method used to determine the association between genetically determined traits, such as gene expression and plasma protein levels, and outcomes of interest, such as disease phenotypes. This method integrates multi-omics data to facilitate the exploration of potential causal relationships between specific drug targets and diseases. To validate the observed causal associations, the HEIDI^96^ test is applied; a p-value below 0.01 indicates the presence of linkage disequilibrium and pleiotropy. For sensitivity analysis, SMR employs multi-SNP-SMR for each circulating protein, setting a significance threshold at p < 0.05, strengthening the main analysis’s evidence. The significance level is adjusted for multiple testing using the Bonferroni method, establishing the threshold at *p* < 5.23 × 10^-7^ (0.05 divided by 2,730 unique proteins and 35 traits). We then performed Bayesian colocalization analysis to complement our SMR findings and determine whether the same causal variants underlie the associations between protein levels and disease phenotypes. A posterior probability (PP.H4) greater than 0.7 indicates significant colocalization, suggesting that identical genetic factors may influence protein abundance and disease outcomes concurrently.

To assess the therapeutic potential of the identified proteins, we utilized the OpenTargets database, which compiles comprehensive data from 22 renowned sources. These sources include genetic associations, somatic mutations, existing drugs, differential expression, animal models, and insights into pathways and systems biology, providing a rich foundation for identifying potential therapeutic applications. The database further enhances our analysis by categorizing protein-related drugs according to their clinical trial phases, as reported on the ClinicalTrials website. Additionally, to ensure the reliability of our findings, we cross-referenced the identified proteins against a well-curated list of druggable genes. Finally, we thoroughly examined the drugs associated with these target proteins, assessing their relevance and potential efficacy based on current clinical and preclinical evidence.

## Declaration of interests

The authors declare no competing interests.

## Data Availability

All data produced in the present work are contained in the manuscript.

ftp://ftp.sanger.ac.uk/pub/project/humgen/summary_statistics/UKBB_blood_cell_traits/

https://www.ebi.ac.uk/gwas/studies/GCST006414

https://cvd.hugeamp.org/datasets.html

https://www.decode.com/summarydata/

https://www.ebi.ac.uk/gwas/studies/GCST009541

https://cvd.hugeamp.org/datasets.html

https://www.ebi.ac.uk/gwas/studies/GCST90104539

## Acknowledgements

Not applicable.

## Author contributions

L.K., F.L., B.Z. and Y.L. conceptualized and supervised this project and wrote the manuscript. L.K. and K.Y. performed the main analyses and wrote the manuscript. L.K., L.Z., Y.Z., M.C., and Z.H. performed the statistical analysis and assisted with interpreting the results. Q.W. and Q.F. provided expertise in cardiovascular biology and GWAS summary statistics. All authors provided intellectual content and approved the final version of the manuscript.

## Notes

### Competing Interest Statement

The authors have declared no competing interest.

### Funding Statement

This study did not receive any funding.

### Author Declarations

The study used only openly available GWAS summary statistics on blood cell traits and cardiovascular diseases that have originally been conducted using human data. GWAS summary statistics on BCTs are available at ftp://ftp.sanger.ac.uk/pub/project/humgen/summary_statistics/UKBB_blood_cell_traits/. GWAS summary statistics on AF, HF, and Stroke are available at the GWAS Catalog (GCST90104539, GCST009541, and GCST90104539). GWAS summary statistics on CAD and PAD are publicly available for download at the Cardiovascular Disease Knowledge Portal (CVDKP) website: https://cvd.hugeamp.org/datasets.html. GWAS summary statistics on VTE are obtained from the deCODE genetics website: https://www.decode.com/summarydata/.

## References

1. Roth GA, Forouzanfar MH, Moran AE, Barber R, Nguyen G, Feigin VL, Naghavi M, Mensah GA, Murray CJ. Demographic and epidemiologic drivers of global cardiovascular mortality. N Engl J Med. 2015;372(14):1333–41. doi:10.1056/NEJMoa1406656

2. Monteiro Júnior JGM, de Oliveira Cipriano Torres D, Filho DCS. Hematological Parameters as Prognostic Biomarkers in Patients with Cardiovascular Diseases. Curr Cardiol Rev. 2019;15(4):274–282. doi:10.2174/1573403x15666190225123544

3. Frostegård J. Immunity, atherosclerosis and cardiovascular disease. BMC Med. 2013;11:117. doi:10.1186/1741-7015-11-117

4. Libby P. Inflammation in atherosclerosis. Nature. 2002;420(6917):868–74. doi:10.1038/nature01323

5. Michel JB, Martin-Ventura JL. Red Blood Cells and Hemoglobin in Human Atherosclerosis and Related Arterial Diseases. Int J Mol Sci. 2020;21(18)doi:10.3390/ijms21186756

6. Businaro R, Tagliani A, Buttari B, Profumo E, Ippoliti F, Di Cristofano C, Capoano R, Salvati B, Riganò R. Cellular and molecular players in the atherosclerotic plaque progression. Ann N Y Acad Sci. 2012;1262:134–41. doi:10.1111/j.1749-6632.2012.06600.x

7. Lievens D, von Hundelshausen P. Platelets in atherosclerosis. Thromb Haemost. 2011;106(5):827–38. doi:10.1160/th11-08-0592

8. Salvagno GL, Sanchis-Gomar F, Picanza A, Lippi G. Red blood cell distribution width: A simple parameter with multiple clinical applications. Crit Rev Clin Lab Sci. 2015;52(2):86–105. doi:10.3109/10408363.2014.992064

9. Kannel WB, Anderson K, Wilson PW. White blood cell count and cardiovascular disease. Insights from the Framingham Study. Jama. 1992;267(9):1253–6.

10. Waterhouse DF, Cahill RA, Sheehan F, McCreery C. Prediction of calculated future cardiovascular disease by monocyte count in an asymptomatic population. Vasc Health Risk Manag. 2008;4(1):177–87. doi:10.2147/vhrm.2008.04.01.177

11. Núñez J, Miñana G, Bodí V, Núñez E, Sanchis J, Husser O, Llàcer A. Low lymphocyte count and cardiovascular diseases. Curr Med Chem. 2011;18(21):3226–33. doi:10.2174/092986711796391633

12. Guasti L, Dentali F, Castiglioni L, Maroni L, Marino F, Squizzato A, Ageno W, Gianni M, Gaudio G, Grandi AM, et al. Neutrophils and clinical outcomes in patients with acute coronary syndromes and/or cardiac revascularisation. A systematic review on more than 34,000 subjects. Thromb Haemost. 2011;106(4):591–9. doi:10.1160/th11-02-0096

13. Haybar H, Pezeshki SMS, Saki N. Evaluation of complete blood count parameters in cardiovascular diseases: An early indicator of prognosis? Exp Mol Pathol. 2019;110:104267. doi:10.1016/j.yexmp.2019.104267

14. Williams PT. Quantile-Specific Heritability of Mean Platelet Volume, Leukocyte Count, and Other Blood Cell Phenotypes. Lifestyle Genom. 2022;15(4):111–123. doi:10.1159/000527048

15. Lin BD, Carnero-Montoro E, Bell JT, Boomsma DI, de Geus EJ, Jansen R, Kluft C, Mangino M, Penninx B, Spector TD, et al. 2SNP heritability and effects of genetic variants for neutrophil-to-lymphocyte and platelet-to-lymphocyte ratio. J Hum Genet. 2017;62(11):979–988. doi:10.1038/jhg.2017.76

16. Vuckovic D, Bao EL, Akbari P, Lareau CA, Mousas A, Jiang T, Chen MH, Raffield LM, Tardaguila M, Huffman JE, et al. The Polygenic and Monogenic Basis of Blood Traits and Diseases. Cell. 2020;182(5):1214–1231.e11. doi:10.1016/j.cell.2020.08.008

17. Astle WJ, Elding H, Jiang T, Allen D, Ruklisa D, Mann AL, Mead D, Bouman H, Riveros-Mckay F, Kostadima MA, et al. The Allelic Landscape of Human Blood Cell Trait Variation and Links to Common Complex Disease. Cell. 2016;167(5):1415–1429.e19. doi:10.1016/j.cell.2016.10.042

18. Zhang X, Johnson AD, Hendricks AE, Hwang SJ, Tanriverdi K, Ganesh SK, Smith NL, Peyser PA, Freedman JE, O’Donnell CJ. Genetic associations with expression for genes implicated in GWAS studies for atherosclerotic cardiovascular disease and blood phenotypes. Hum Mol Genet. 2014;23(3):782–95. doi:10.1093/hmg/ddt461

19. Paaby AB, Rockman MV. The many faces of pleiotropy. Trends Genet. 2013;29(2):66–73. doi:10.1016/j.tig.2012.10.010

20. Harshfield EL, Sims MC, Traylor M, Ouwehand WH, Markus HS. The role of haematological traits in risk of ischaemic stroke and its subtypes. Brain. 2020;143(1):210–221. doi:10.1093/brain/awz362

21. He J, Jiang Q, Yao Y, Shen Y, Li J, Yang J, Ma R, Zhang N, Liu C. Blood Cells and Venous Thromboembolism Risk: A Two-Sample Mendelian Randomization Study. Front Cardiovasc Med. 2022;9:919640. doi:10.3389/fcvm.2022.919640

22. Yang Y, Zhou Y, Nyholt DR, Yap CX, Tannenberg RK, Wang Y, Wu Y, Zhu Z, Taylor BV, Gratten J. The shared genetic landscape of blood cell traits and risk of neurological and psychiatric disorders. Cell Genom. 2023;3(2):100249. doi:10.1016/j.xgen.2022.100249

23. den Harder AM, de Jong PA, de Groot MCH, Wolterink JM, Budde RPJ, Iŝgum I, van Solinge WW, Ten Berg MJ, Lutgens E, Veldhuis WB, et al. Commonly available hematological biomarkers are associated with the extent of coronary calcifications. Atherosclerosis. 2018;275:166–173. doi:10.1016/j.atherosclerosis.2018.06.017

24. Nikpay M, Mohammadzadeh S. Phenome-wide screening for traits causally associated with the risk of coronary artery disease. J Hum Genet. 2020;65(4):371–380. doi:10.1038/s10038-019-0716-z

25. Ellingsen TS, Lappegård J, Skjelbakken T, Brækkan SK, Hansen JB. Red cell distribution width is associated with incident venous thromboembolism (VTE) and case-fatality after VTE in a general population. Thromb Haemost. 2015;113(1):193–200. doi:10.1160/th14-04-0335

26. Bucciarelli P, Maino A, Felicetta I, Abbattista M, Passamonti SM, Artoni A, Martinelli I. Association between red cell distribution width and risk of venous thromboembolism. Thromb Res. 2015;136(3):590–4. doi:10.1016/j.thromres.2015.07.020

27. Varga-Szabo D, Pleines I, Nieswandt B. Cell adhesion mechanisms in platelets. Arterioscler Thromb Vasc Biol. 2008;28(3):403–12. doi:10.1161/atvbaha.107.150474

28. Hansen M, Zeddies S, Meinders M, di Summa F, van Alphen FPJ, Hoogendijk AJ, Moore KS, Halbach M, Gutiérrez L, van den Biggelaar M, et al. The RNA-Binding Protein ATXN2 is Expressed during Megakaryopoiesis and May Control Timing of Gene Expression. Int J Mol Sci. 2020;21(3)doi:10.3390/ijms21030967

29. Nemkov T, Stephenson D, Erickson C, Dzieciatkowska M, Key A, Moore A, Earley EJ, Page GP, Lacroix IS, Stone M, et al. Regulation of kynurenine metabolism by blood donor genetics and biology impacts red cell hemolysis in vitro and in vivo. Blood. 2024;143(5):456–472. doi:10.1182/blood.2023022052

30. Ganesh SK, Tragante V, Guo W, Guo Y, Lanktree MB, Smith EN, Johnson T, Castillo BA, Barnard J, Baumert J, et al. Loci influencing blood pressure identified using a cardiovascular gene-centric array. Hum Mol Genet. 2013;22(8):1663–78. doi:10.1093/hmg/dds555

31. Paterson AD, Lopes-Virella MF, Waggott D, Boright AP, Hosseini SM, Carter RE, Shen E, Mirea L, Bharaj B, Sun L, et al. Genome-wide association identifies the ABO blood group as a major locus associated with serum levels of soluble E-selectin. Arterioscler Thromb Vasc Biol. 2009;29(11):1958–67. doi:10.1161/atvbaha.109.192971

32. Prugger C, Luc G, Haas B, Morange PE, Ferrieres J, Amouyel P, Kee F, Ducimetiere P, Empana JP. Multiple biomarkers for the prediction of ischemic stroke: the PRIME study. Arterioscler Thromb Vasc Biol. 2013;33(3):659–66. doi:10.1161/atvbaha.112.300109

33. Mendez-Barbero N, Yuste-Montalvo A, Nuñez-Borque E, Jensen BM, Gutiérrez-Muñoz C, Tome-Amat J, Garrido-Arandia M, Díaz-Perales A, Ballesteros-Martinez C, Laguna JJ, et al. The TNF-like weak inducer of the apoptosis/fibroblast growth factor-inducible molecule 14 axis mediates histamine and platelet-activating factor-induced subcutaneous vascular leakage and anaphylactic shock. J Allergy Clin Immunol. 2020;145(2):583–596.e6. doi:10.1016/j.jaci.2019.09.019

34. Franchini M, Lippi G. Relative Risks of Thrombosis and Bleeding in Different ABO Blood Groups. Semin Thromb Hemost. 2016;42(2):112–7. doi:10.1055/s-0035-1564832

35. Lee JY, Lee BS, Shin DJ, Woo Park K, Shin YA, Joong Kim K, Heo L, Young Lee J, Kyoung Kim Y, Jin Kim Y, et al. A genome-wide association study of a coronary artery disease risk variant. J Hum Genet. 2013;58(3):120–6. doi:10.1038/jhg.2012.124

36. Méndez-Barbero N, Gutiérrez-Muñoz C, Blázquez-Serra R, Martín-Ventura JL, Blanco-Colio LM. Tumor Necrosis Factor-Like Weak Inducer of Apoptosis (TWEAK)/Fibroblast Growth Factor-Inducible 14 (Fn14) Axis in Cardiovascular Diseases: Progress and Challenges. Cells. 2020;9(2)doi:10.3390/cells9020405

37. Nagai M, Hirayama K, Ebihara I, Higuchi T, Imaizumi M, Maruyama H, Miyamoto Y, Kakita T, Ogawa Y, Fujita S, et al. Serum TNF-related and weak inducer of apoptosis levels in septic shock patients. Ther Apher Dial. 2011;15(4):342–8. doi:10.1111/j.1744-9987.2011.00966.x

38. Chasman DI, Paré G, Mora S, Hopewell JC, Peloso G, Clarke R, Cupples LA, Hamsten A, Kathiresan S, Mälarstig A, et al. Forty-three loci associated with plasma lipoprotein size, concentration, and cholesterol content in genome-wide analysis. PLoS Genet. 2009;5(11):e1000730. doi:10.1371/journal.pgen.1000730

39. Ren L, Du W, Song D, Lu H, Hamblin MH, Wang C, Du C, Fan GC, Becker RC, Fan Y. Genetic ablation of diabetes-associated gene Ccdc92 reduces obesity and insulin resistance in mice. iScience. 2023;26(1):105769. doi:10.1016/j.isci.2022.105769

40. Ellingsen TS, Lappegård J, Ueland T, Aukrust P, Brækkan SK, Hansen JB. Plasma hepcidin is associated with future risk of venous thromboembolism. Blood Adv. 2018;2(11):1191–1197. doi:10.1182/bloodadvances.2018018465

41. Luo J, Thomassen JQ, Nordestgaard BG, Tybjærg-Hansen A, Frikke-Schmidt R. Neutrophil counts and cardiovascular disease. Eur Heart J. 2023;44(47):4953–4964. doi:10.1093/eurheartj/ehad649

42. Weymann A, Ali-Hasan-Al-Saegh S, Sabashnikov A, Popov AF, Mirhosseini SJ, Nombela-Franco L, Testa L, Lotfaliani M, Zeriouh M, Liu T, et al. Platelets Cellular and Functional Characteristics in Patients with Atrial Fibrillation: A Comprehensive Meta-Analysis and Systematic Review. Med Sci Monit Basic Res. 2017;23:58–86. doi:10.12659/msmbr.902557

43. Kim YK, Hwang MY, Kim YJ, Moon S, Han S, Kim BJ. Evaluation of pleiotropic effects among common genetic loci identified for cardio-metabolic traits in a Korean population. Cardiovasc Diabetol. 2016;15:20. doi:10.1186/s12933-016-0337-1

44. Kato N, Takeuchi F, Tabara Y, Kelly TN, Go MJ, Sim X, Tay WT, Chen CH, Zhang Y, Yamamoto K, et al. Meta-analysis of genome-wide association studies identifies common variants associated with blood pressure variation in east Asians. Nat Genet. 2011;43(6):531–8. doi:10.1038/ng.834

45. Zhang J, Guo Y, Zhao X, Pang J, Pan C, Wang J, Wei S, Yu X, Zhang C, Chen Y, et al. The role of aldehyde dehydrogenase 2 in cardiovascular disease. Nat Rev Cardiol. 2023;20(7):495–509. doi:10.1038/s41569-023-00839-5

46. Wang W, Tang Y, Wang Y, Tascau L, Balcerek J, Tong W, Levine RL, Welch C, Tall AR, Wang N. LNK/SH2B3 Loss of Function Promotes Atherosclerosis and Thrombosis. Circ Res. 2016;119(6):e91–e103. doi:10.1161/circresaha.116.308955

47. Bersenev A, Wu C, Balcerek J, Tong W. Lnk controls mouse hematopoietic stem cell self-renewal and quiescence through direct interactions with JAK2. J Clin Invest. 2008;118(8):2832–44. doi:10.1172/jci35808

48. Deloukas P, Kanoni S, Willenborg C, Farrall M, Assimes TL, Thompson JR, Ingelsson E, Saleheen D, Erdmann J, Goldstein BA, et al. Large-scale association analysis identifies new risk loci for coronary artery disease. Nat Genet. 2013;45(1):25–33. doi:10.1038/ng.2480

49. Sahadevan P, Allen BG. MK5: A novel regulator of cardiac fibroblast function? IUBMB Life. 2017;69(10):785–794. doi:10.1002/iub.1677

50. Yoshizuka N, Chen RM, Xu Z, Liao R, Hong L, Hu WY, Yu G, Han J, Chen L, Sun P. A novel function of p38-regulated/activated kinase in endothelial cell migration and tumor angiogenesis. Mol Cell Biol. 2012;32(3):606–18. doi:10.1128/mcb.06301-11

51. Page GP, Kanias T, Guo YJ, Lanteri MC, Zhang X, Mast AE, Cable RG, Spencer BR, Kiss JE, Fang F, et al. Multiple-ancestry genome-wide association study identifies 27 loci associated with measures of hemolysis following blood storage. J Clin Invest. 2021;131(13)doi:10.1172/jci146077

52. Erbilgin A, Civelek M, Romanoski CE, Pan C, Hagopian R, Berliner JA, Lusis AJ. Identification of CAD candidate genes in GWAS loci and their expression in vascular cells. J Lipid Res. 2013;54(7):1894–905. doi:10.1194/jlr.M037085

53. He M, Pei Z, Mohsen AW, Watkins P, Murdoch G, Van Veldhoven PP, Ensenauer R, Vockley J. Identification and characterization of new long chain acyl-CoA dehydrogenases. Mol Genet Metab. 2011;102(4):418–29. doi:10.1016/j.ymgme.2010.12.005

54. Xiao S, Qi M, Zhou Q, Gong H, Wei D, Wang G, Feng Q, Wang Z, Liu Z, Zhou Y, et al. Macrophage fatty acid oxidation in atherosclerosis. Biomed Pharmacother. 2024;170:116092. doi:10.1016/j.biopha.2023.116092

55. Leppänen O, Björnheden T, Evaldsson M, Borén J, Wiklund O, Levin M. ATP depletion in macrophages in the core of advanced rabbit atherosclerotic plaques in vivo. Atherosclerosis. 2006;188(2):323–30. doi:10.1016/j.atherosclerosis.2005.11.017

56. Hao L, Ren M, Rong B, Xie F, Lin MJ, Zhao YC, Yue X, Han WQ, Zhong JQ. TWEAK/Fn14 mediates atrial-derived HL-1 myocytes hypertrophy via JAK2/STAT3 signalling pathway. J Cell Mol Med. 2018;22(9):4344–4353. doi:10.1111/jcmm.13724

57. Inoue T, Matsunaga R, Sakai Y, Yaguchi I, Takayanagi K, Morooka S. Insulin resistance affects endothelium-dependent acetylcholine-induced coronary artery response. Eur Heart J. 2000;21(11):895–900. doi:10.1053/euhj.1999.1872

58. Neville MJ, Wittemans LBL, Pinnick KE, Todorčević M, Kaksonen R, Pietiläinen KH, Luan J, Scott RA, Wareham NJ, Langenberg C, et al. Regional fat depot masses are influenced by protein-coding gene variants. PLoS One. 2019;14(5):e0217644. doi:10.1371/journal.pone.0217644

59. Kraja AT, Chasman DI, North KE, Reiner AP, Yanek LR, Kilpeläinen TO, Smith JA, Dehghan A, Dupuis J, Johnson AD, et al. Pleiotropic genes for metabolic syndrome and inflammation. Mol Genet Metab. 2014;112(4):317–38. doi:10.1016/j.ymgme.2014.04.007

60. Xu S, Ilyas I, Little PJ, Li H, Kamato D, Zheng X, Luo S, Li Z, Liu P, Han J, et al. Endothelial Dysfunction in Atherosclerotic Cardiovascular Diseases and Beyond: From Mechanism to Pharmacotherapies. Pharmacol Rev. 2021;73(3):924–967. doi:10.1124/pharmrev.120.000096

61. Woollard KJ, Geissmann F. Monocytes in atherosclerosis: subsets and functions. Nat Rev Cardiol. 2010;7(2):77–86. doi:10.1038/nrcardio.2009.228

62. Cyr Y, Bozal FK, Barcia Durán JG, Newman AAC, Amadori L, Smyrnis P, Gourvest M, Das D, Gildea M, Kaur R, et al. The IRG1-itaconate axis protects from cholesterol-induced inflammation and atherosclerosis. Proc Natl Acad Sci U S A. 2024;121(15):e2400675121. doi:10.1073/pnas.2400675121

63. Fuentes F, Palomo I, Fuentes E. Platelet oxidative stress as a novel target of cardiovascular risk in frail older people. Vascul Pharmacol. 2017;93-95:14–19. doi:10.1016/j.vph.2017.07.003

64. Montenont E, Echagarruga C, Allen N, Araldi E, Suarez Y, Berger JS. Platelet WDR1 suppresses platelet activity and is associated with cardiovascular disease. Blood. 2016;128(16):2033–2042. doi:10.1182/blood-2016-03-703157

65. Gleissner CA. Platelet-derived chemokines in atherogenesis: what’s new? Curr Vasc Pharmacol. 2012;10(5):563–9. doi:10.2174/157016112801784521

66. Zahid M, Mangin P, Loyau S, Hechler B, Billiald P, Gachet C, Jandrot-Perrus M. The future of glycoprotein VI as an antithrombotic target. J Thromb Haemost. 2012;10(12):2418–27. doi:10.1111/jth.12009

67. Jeney V, Balla G, Balla J. Red blood cell, hemoglobin and heme in the progression of atherosclerosis. Front Physiol. 2014;5:379. doi:10.3389/fphys.2014.00379

68. Clemetson KJ, Clemetson JM. Platelet collagen receptors. Thromb Haemost. 2001;86(1):189–97.

69. Nielsen JB, Thorolfsdottir RB, Fritsche LG, Zhou W, Skov MW, Graham SE, Herron TJ, McCarthy S, Schmidt EM, Sveinbjornsson G, et al. Biobank-driven genomic discovery yields new insight into atrial fibrillation biology. Nat Genet. 2018;50(9):1234–1239. doi:10.1038/s41588-018-0171-3

70. Aragam KG, Jiang T, Goel A, Kanoni S, Wolford BN, Atri DS, Weeks EM, Wang M, Hindy G, Zhou W, et al. Discovery and systematic characterization of risk variants and genes for coronary artery disease in over a million participants. Nat Genet. 2022;54(12):1803–1815. doi:10.1038/s41588-022-01233-6

71. Ghouse J, Tragante V, Ahlberg G, Rand SA, Jespersen JB, Leinøe EB, Vissing CR, Trudsø L, Jonsdottir I, Banasik K, et al. Genome-wide meta-analysis identifies 93 risk loci and enables risk prediction equivalent to monogenic forms of venous thromboembolism. Nat Genet. 2023;55(3):399–409. doi:10.1038/s41588-022-01286-7

72. Shah S, Henry A, Roselli C, Lin H, Sveinbjörnsson G, Fatemifar G, Hedman Å K, Wilk JB, Morley MP, Chaffin MD, et al. Genome-wide association and Mendelian randomisation analysis provide insights into the pathogenesis of heart failure. Nat Commun. 2020;11(1):163. doi:10.1038/s41467-019-13690-5

73. van Zuydam NR, Stiby A, Abdalla M, Austin E, Dahlström EH, McLachlan S, Vlachopoulou E, Ahlqvist E, Di Liao C, Sandholm N, et al. Genome-Wide Association Study of Peripheral Artery Disease. Circ Genom Precis Med. 2021;14(5):e002862. doi:10.1161/circgen.119.002862

74. Mishra A, Malik R, Hachiya T, Jürgenson T, Namba S, Posner DC, Kamanu FK, Koido M, Le Grand Q, Shi M, et al. Stroke genetics informs drug discovery and risk prediction across ancestries. Nature. 2022;611(7934):115–123. doi:10.1038/s41586-022-05165-3

75. Tobin MD, Minelli C, Burton PR, Thompson JR. Commentary: development of Mendelian randomization: from hypothesis test to ’Mendelian deconfounding’. Int J Epidemiol. 2004;33(1):26–9. doi:10.1093/ije/dyh016

76. Auton A, Brooks LD, Durbin RM, Garrison EP, Kang HM, Korbel JO, Marchini JL, McCarthy S, McVean GA, Abecasis GR. A global reference for human genetic variation. Nature. 2015;526(7571):68–74. doi:10.1038/nature15393

77. Finucane HK, Reshef YA, Anttila V, Slowikowski K, Gusev A, Byrnes A, Gazal S, Loh PR, Lareau C, Shoresh N, et al. Heritability enrichment of specifically expressed genes identifies disease-relevant tissues and cell types. Nat Genet. 2018;50(4):621–629. doi:10.1038/s41588-018-0081-4

78. Human genomics. The Genotype-Tissue Expression (GTEx) pilot analysis: multitissue gene regulation in humans. Science. 2015;348(6235):648–60. doi:10.1126/science.1262110

79. Foley CN, Staley JR, Breen PG, Sun BB, Kirk PDW, Burgess S, Howson JMM. A fast and efficient colocalization algorithm for identifying shared genetic risk factors across multiple traits. Nat Commun. 2021;12(1):764. doi:10.1038/s41467-020-20885-8

80. Duncan LE, Shen H, Ballon JS, Hardy KV, Noordsy DL, Levinson DF. Genetic Correlation Profile of Schizophrenia Mirrors Epidemiological Results and Suggests Link Between Polygenic and Rare Variant (22q11.2) Cases of Schizophrenia. Schizophr Bull. 2018;44(6):1350–1361. doi:10.1093/schbul/sbx174

81. Chung D, Yang C, Li C, Gelernter J, Zhao H. GPA: a statistical approach to prioritizing GWAS results by integrating pleiotropy and annotation. PLoS Genet. 2014;10(11):e1004787. doi:10.1371/journal.pgen.1004787

82. Purcell S, Neale B, Todd-Brown K, Thomas L, Ferreira MA, Bender D, Maller J, Sklar P, de Bakker PI, Daly MJ, et al. PLINK: a tool set for whole-genome association and population-based linkage analyses. Am J Hum Genet. 2007;81(3):559–75. doi:10.1086/519795

83. Darrous L, Mounier N, Kutalik Z. Simultaneous estimation of bi-directional causal effects and heritable confounding from GWAS summary statistics. Nat Commun. 2021;12(1):7274. doi:10.1038/s41467-021-26970-w

84. Burgess S, Bowden J, Fall T, Ingelsson E, Thompson SG. Sensitivity Analyses for Robust Causal Inference from Mendelian Randomization Analyses with Multiple Genetic Variants. Epidemiology. 2017;28(1):30–42. doi:10.1097/ede.0000000000000559

85. Ray D, Chatterjee N. A powerful method for pleiotropic analysis under composite null hypothesis identifies novel shared loci between Type 2 Diabetes and Prostate Cancer. PLoS Genet. 2020;16(12):e1009218. doi:10.1371/journal.pgen.1009218

86. Watanabe K, Taskesen E, van Bochoven A, Posthuma D. Functional mapping and annotation of genetic associations with FUMA. Nat Commun. 2017;8(1):1826. doi:10.1038/s41467-017-01261-5

87. Kircher M, Witten DM, Jain P, O’Roak BJ, Cooper GM, Shendure J. A general framework for estimating the relative pathogenicity of human genetic variants. Nat Genet. 2014;46(3):310–5. doi:10.1038/ng.2892

88. Boyle AP, Hong EL, Hariharan M, Cheng Y, Schaub MA, Kasowski M, Karczewski KJ, Park J, Hitz BC, Weng S, et al. Annotation of functional variation in personal genomes using RegulomeDB. Genome Res. 2012;22(9):1790–7. doi:10.1101/gr.137323.112

89. Kundaje A, Meuleman W, Ernst J, Bilenky M, Yen A, Heravi-Moussavi A, Kheradpour P, Zhang Z, Wang J, Ziller MJ, et al. Integrative analysis of 111 reference human epigenomes. Nature. 2015;518(7539):317–30. doi:10.1038/nature14248

90. Giambartolomei C, Vukcevic D, Schadt EE, Franke L, Hingorani AD, Wallace C, Plagnol V. Bayesian test for colocalisation between pairs of genetic association studies using summary statistics. PLoS Genet. 2014;10(5):e1004383. doi:10.1371/journal.pgen.1004383

91. Wallace C. Eliciting priors and relaxing the single causal variant assumption in colocalisation analyses. PLoS Genet. 2020;16(4):e1008720. doi:10.1371/journal.pgen.1008720

92. Gerring ZF, Mina-Vargas A, Gamazon ER, Derks EM. E-MAGMA: an eQTL-informed method to identify risk genes using genome-wide association study summary statistics. Bioinformatics. 2021;37(16):2245–2249. doi:10.1093/bioinformatics/btab115

93. Zhu H, Zhou X. Transcriptome-wide association studies: a view from Mendelian randomization. Quant Biol. 2021;9(2):107–121. doi:10.1007/s40484-020-0207-4

94. de Leeuw CA, Mooij JM, Heskes T, Posthuma D. MAGMA: generalized gene-set analysis of GWAS data. PLoS Comput Biol. 2015;11(4):e1004219. doi:10.1371/journal.pcbi.1004219

95. Zhou Y, Zhou B, Pache L, Chang M, Khodabakhshi AH, Tanaseichuk O, Benner C, Chanda SK. Metascape provides a biologist-oriented resource for the analysis of systems-level datasets. Nat Commun. 2019;10(1):1523. doi:10.1038/s41467-019-09234-6

96. Zhu Z, Zhang F, Hu H, Bakshi A, Robinson MR, Powell JE, Montgomery GW, Goddard ME, Wray NR, Visscher PM, et al. Integration of summary data from GWAS and eQTL studies predicts complex trait gene targets. Nat Genet. 2016;48(5):481–7. doi:10.1038/ng.3538

