## Supplementary Information for "Unveiling Shared Genetic Links between Blood Cell Traits and Cardiovascular Diseases"

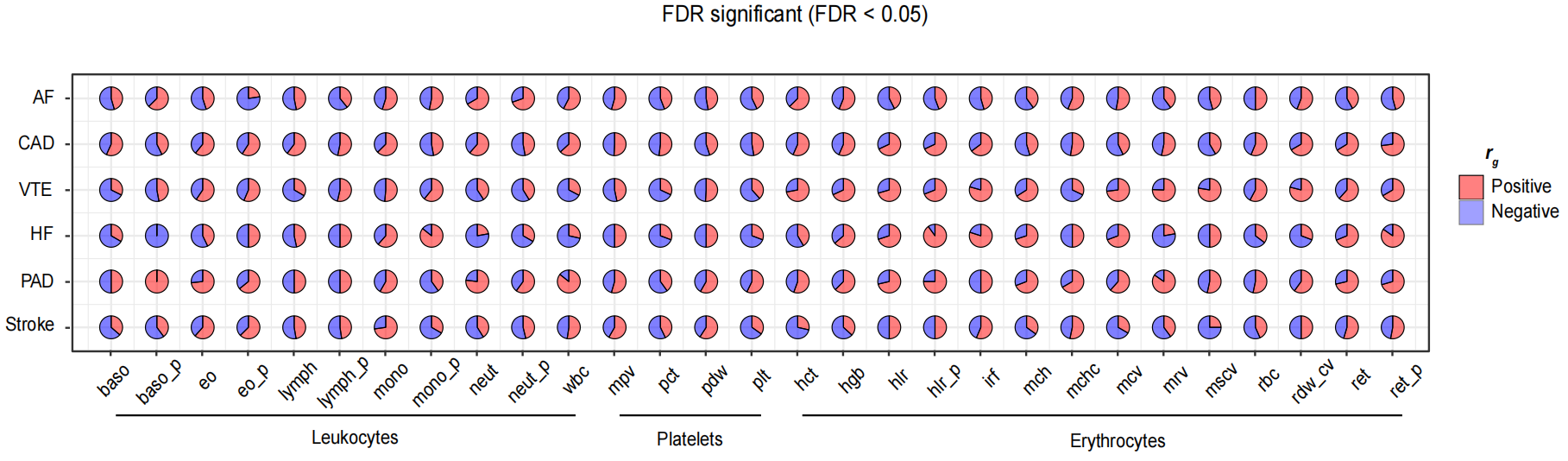
**SFigure 1: Local genetic correlations between BCTs and CVDs estimated using LAVA**

Regions with significant local genetic correlation among the 174 specific trait pairs at the level of FDR correction threshold are shown in pink and vice versa in purple.

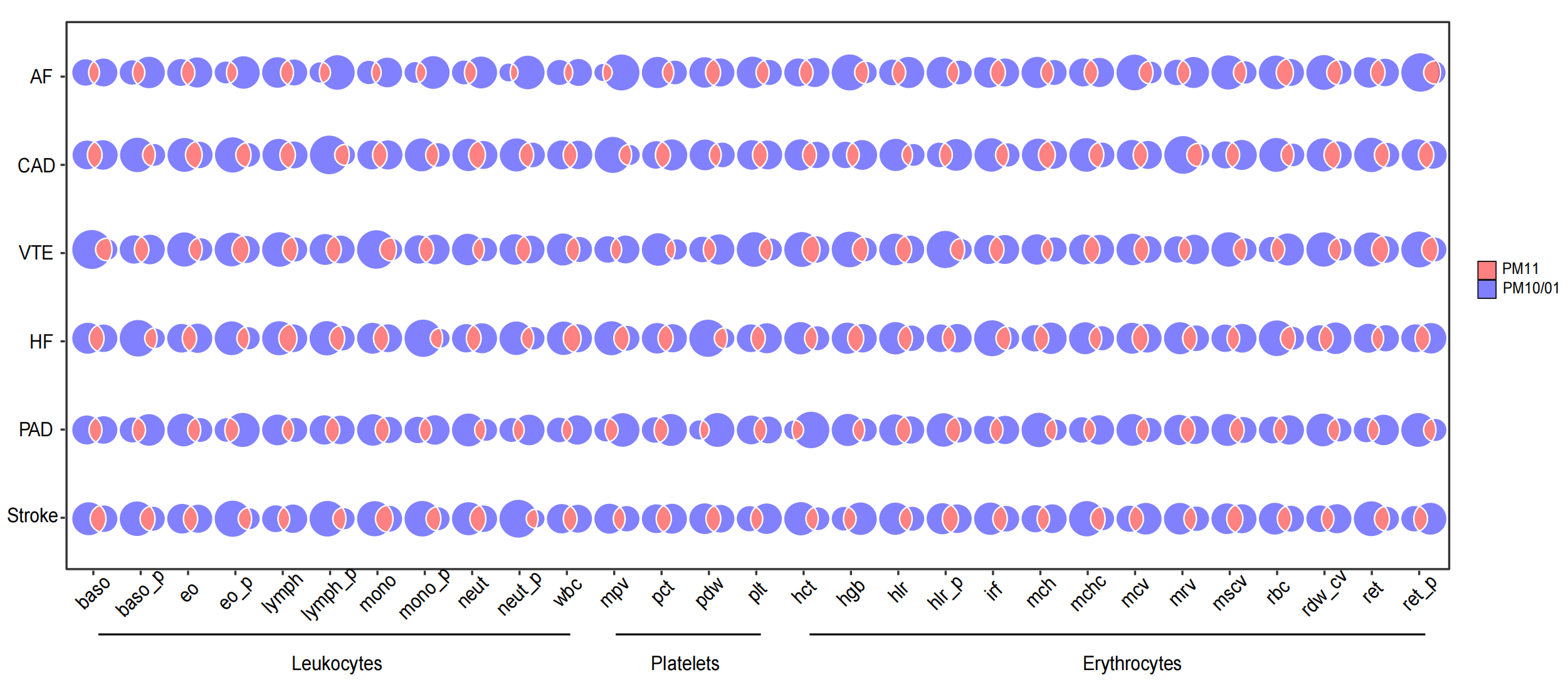

**SFigure 2: Estimated genome-wide genetic overlap between BCTs and CVDs using GPA**

Genetic overlap between BCTs and CVDs. The pink color in the Venn diagram represents the overlap of SNPs shared between BCTs and CVDs, and the purple color represents the unique SNPs of BCTs or CVDs. The size of the circles reflects the degree of effective variants.

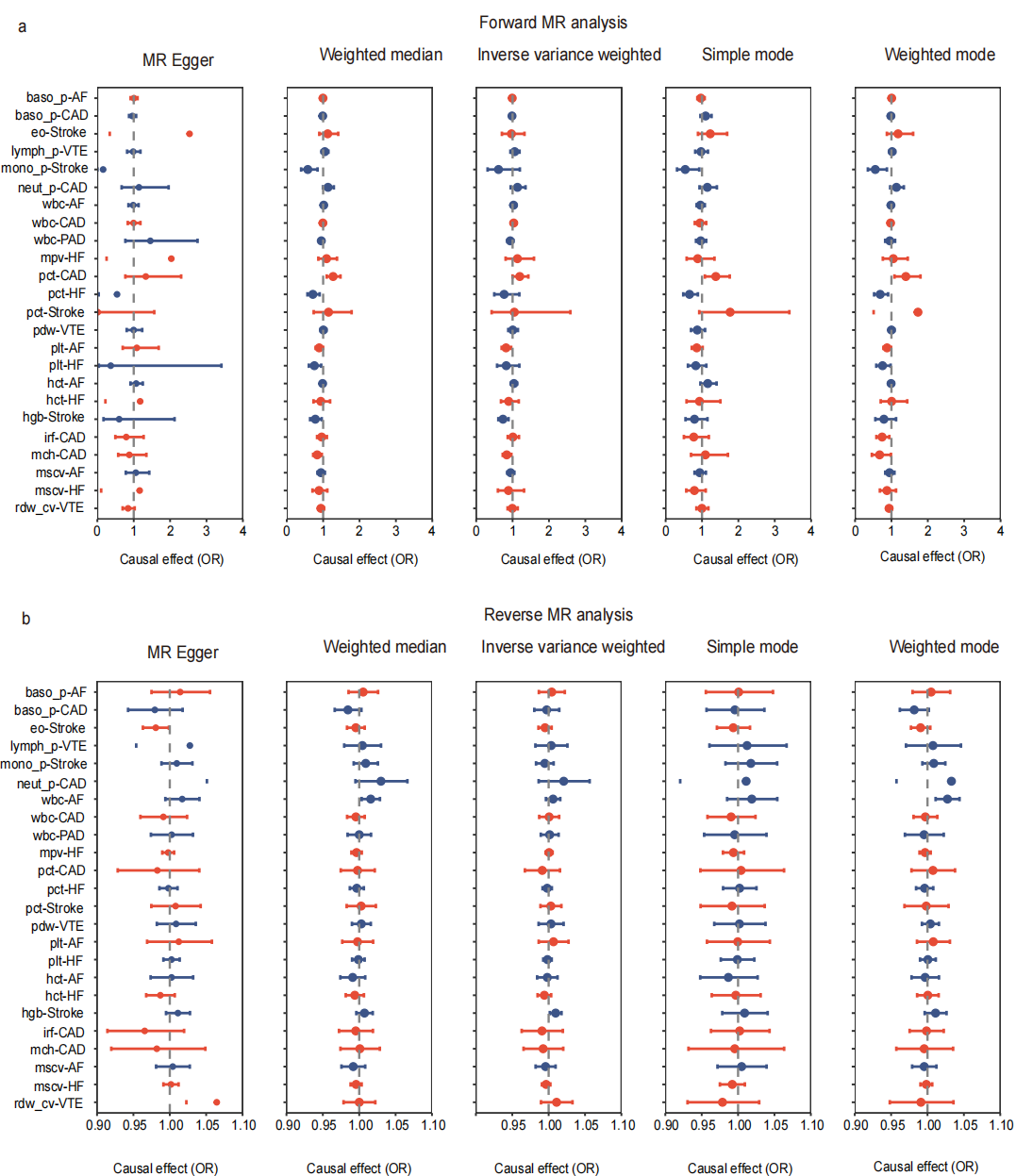

**SFigure 3: Tissue type-specific associations with BCTs and CVDs identified using gene expression data**

Estimating associations between tissue-specific gene expression (GTEx) and BCTs and CVDs using LDSC-SEG. Multiple testing was performed on the 49 tissues, and only those with significant associations (FDR < 0.05) were shown.

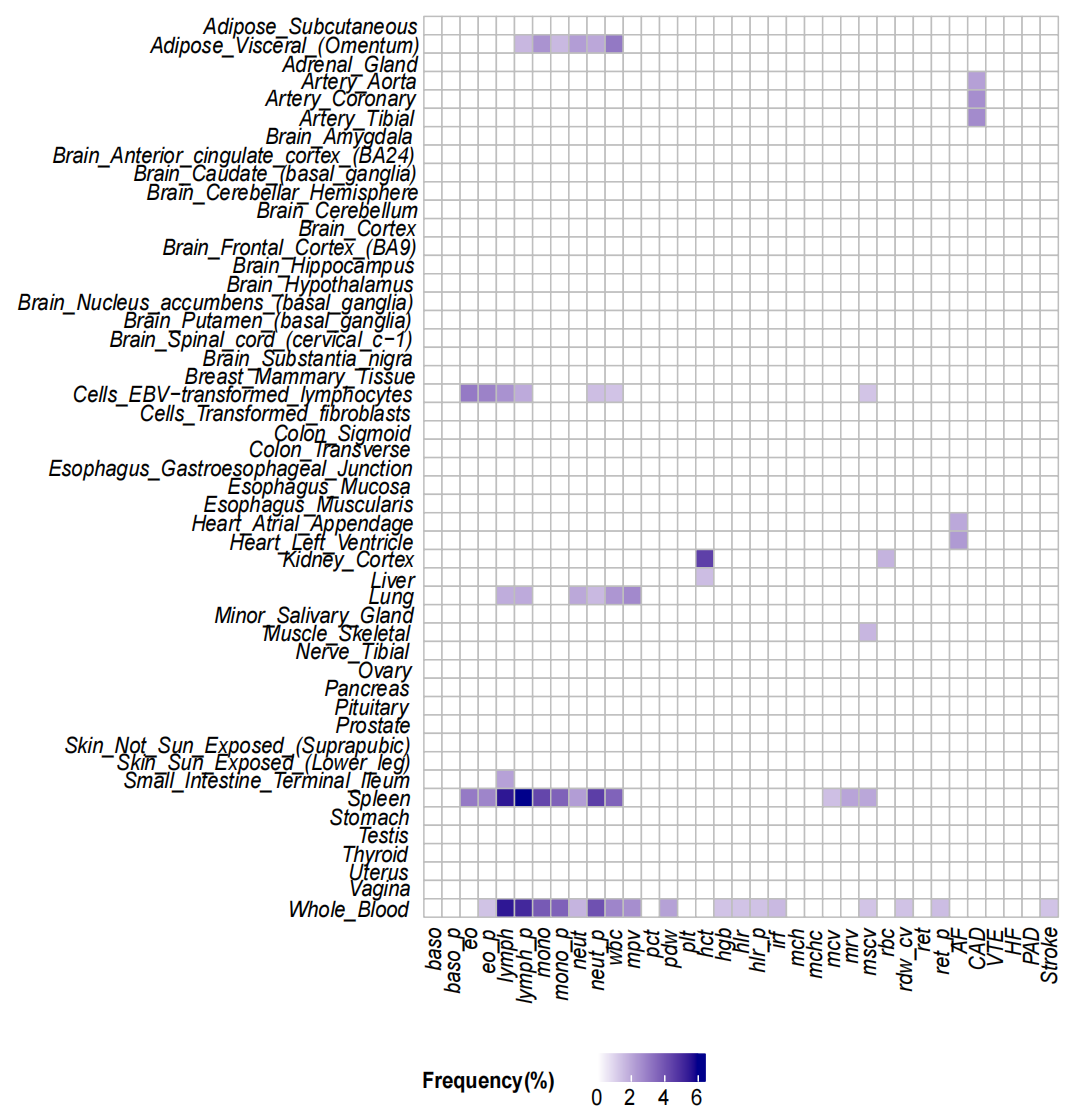

**SFigure 4: Sensitivity analysis of the bidirectional causal relationship between BCTs and CVDs**

Sensitivity analysis was performed using five traditional MR models to validate our findings, including IVW, MR-Egger, weighted median, simple models, and weighted model methods. (a) Sensitivity analysis of forward MR: BCTs as exposure and CVDs as outcome; (b) Sensitivity analysis of reverse MR: CVDs as exposure and BCTs as outcome. Solid dots represent the mean odds ratio, and error bars reflect the 95% confidence intervals. P-values < 0.05 are considered statistically significant. Vertical dashed line set at odds ratio of 1.0.
